# Fasting plasma fructose is associated with postprandial ethanol levels and mitochondrial dysfunction: linking fasting fructose to MASLD

**DOI:** 10.64898/2026.10.30.26366906

**Authors:** Florine H.M. Westerbeke, Ilias Attaye, Abraham S. Meijnikman, Núria Farràs Solé, Artemiy Kovynev, Oluwatomisono I. Akinrimisi, Joanne Verheij, Michail Doukas, Arnold van de Laar, Rutger Franken, Melany Rios-Morales, Max Nieuwdorp

**Affiliations:** Department of Internal and Vascular Medicine, Amsterdam University Medical Center, Amsterdam, the Netherlands; Department of Internal Medicine, Division of Nephrology and Transplantation, Erasmus University Medical Center, Rotterdam, the Netherlands; Department of Gastroenterology and Hepatology, Amsterdam University Medical Center, Amsterdam, the Netherlands; Tytgat Institute for Liver and Intestinal Research, Amsterdam University Medical Center, Amsterdam, the Netherlands; Department of Pathology, Amsterdam University Medical Center, Amsterdam, the Netherlands; Department of Pathology, Antwerp University Hospital / University of Antwerp, Antwerp, Belgium; Department of Pathology, Erasmus University Medical Center, Rotterdam, the Netherlands; Department of Surgery, Spaarne Gasthuis, Hoofddorp, the Netherlands

**Author notes:** **Corresponding author:** Prof. dr. Max Nieuwdorp, dr. Melany Rios-Morales. These authors contributed equally to this work (Melany Rios-Morales, Max Nieuwdorp).

**Keywords:** Fructose, MASLD, gut microbiota, fecal microbiota, mitochondria

## Abstract

High dietary fructose consumption is a recognized risk factor for metabolic dysfunction-associated steatotic liver disease (MASLD), but the clinical relevance of fasting fructose levels remains unexplored. This study investigated gut microbiota and hepatic transcriptional alterations associated with fasting fructose levels and their potential link to MASLD. We analyzed data from a subset of 49 morbidly obese individuals enrolled in the prospective BARIA cohort. Fasting plasma fructose levels were significantly higher in individuals with MASLD and were associated with polyol pathway downregulation and lipogenic gene upregulation. Fasting fructose also positively correlated with postprandial plasma ethanol increase, suggesting enhanced microbial endogenous production and/or altered ethanol catabolism, the latter supported by suppressed hepatic alcohol dehydrogenase gene expression. In addition, fasting fructose was associated with fecal gut microbiota alterations, including significant shifts in beta diversity and correlations with specific bacterial species. At the transcriptomic level, elevated fasting fructose was linked to impaired mitochondrial energy metabolism and antioxidant defense, signatures associated with MASLD-related metabolic disruption. In conclusion, our data indicate that fasting plasma fructose reflects dietary intake rather than endogenous production and increased levels are associated with metabolic and microbiota changes linked to MASLD. Although not intended to replace established glycemic markers, our findings suggest that fasting fructose may provide complementary insights into the metabolic state of the liver and MASLD-related dysregulation, supporting its potential as a MASLD risk biomarker.

## Introduction

Fructose consumption has increased in recent decades, driven by the growing intake of processed foods and beverages containing high amounts of added sugars such as high fructose corn syrup.^1,2^ This increased fructose consumption has been identified as an important risk factor for the development of metabolic dysfunction-associated steatotic liver disease (MASLD),^3–6^ the most prevalent chronic liver disease worldwide.^7^ MASLD not only increases liver-related complications, but also the risk of cardiovascular-and chronic kidney diseases.^8^ An estimated rise of the MASLD prevalence to more than 55% by 2040^9^ highlights the urgent need for novel preventative and therapeutic strategies.

Excessive dietary fructose intake is recognized as an important driver of MASLD due to its unique metabolic effects. In contrast to glycolysis, hepatic fructose catabolism is an unregulated process as it bypasses the rate-limiting steps of glycolysis. The unregulated catabolism of fructose promotes *de novo* lipogenesis (DNL), resulting in excessive production and accumulation of fatty acids (FA) in the liver.^3,10,11^ Moreover, the rapid phosphorylation of fructose in the liver can cause ATP depletion and subsequent uric acid production,^12–14^ which induces oxidative stress and further promotes DNL and insulin resistance.^15–19^ In addition to these direct hepatic metabolic effects, the small intestine has also been suggested to play a crucial direct role (next to dietary uptake) in fructose catabolism.^20^ In murine models, it has been demonstrated that the small intestine can convert physiological doses of dietary fructose into glucose and organic acids.^20^ However, this small intestinal gluconeogenesis was saturated at high fructose doses, leading to fructose overflow to both the liver and the distal parts of the intestine, where it becomes available for gut microbial fermentation. In the colon for example, gut microbiota-derived acetate can be produced from dietary fructose, which can stimulate DNL.^21^ Furthermore, various intestinal microbiota species have been shown to produce ethanol, another driver of DNL, from the fermentation of sugars, including fructose.^22,23^ This gut microbiota derived endogenous ethanol production has been associated with the development of MASLD,^24^ highlighting the potential of the gut-liver axis to contribute to fructose-induced liver disease.

While associations between dietary fructose intake and MASLD pathogenesis are well-established, the significance of fasting plasma fructose concentrations remains underexplored. We used plasma metabolites, liver histology, fecal shotgun metagenomics and liver RNA-sequencing (RNA-seq) data to investigate the associations between fasting plasma fructose levels, the gut microbiota and hepatic gene expression, as well as their potential association with MASLD histology in a morbidly obese population. Using this integrative analysis, we aimed to provide novel insights into the metabolic and microbial signatures associated with fasting plasma fructose levels and their potential role in MASLD pathogenesis. By this means, we investigate whether fasting plasma fructose concentrations may serve as a potential metabolic marker in the context of MASLD.

## Materials and methods

### Study design

For this study a subset of 49 individuals was included from the ongoing BARIA study, a longitudinal cohort study of morbidly obese subjects scheduled for bariatric surgery.^25^ The inclusion criteria for this study were adults, aged <65 years, scheduled for bariatric surgery, with a BMI ≥40 kg/m^2^ or a BMI ≥35 kg/m^2^ with obesity-related comorbidity. Baseline measurements, collected within 3 months before surgery, included demographics, biometrics, fasting blood samples, urine and faecal samples and a two-hour oral mixed meal test (MMT). On the day of the bariatric surgery, preoperative fecal samples and fasting blood samples were collected. Furthermore, tissue biopsies from liver, jejunum and adipose tissues were obtained during surgery. All samples were stored at -80°C until further use. All subjects provided written informed consent and the study is executed in accordance with the Declaration of Helsinki. The study was registered at the Dutch trial registry at 29-03-2016, registration number: NL-OMON50702, and approved by the Academic Medical Center Ethics Committee of the Amsterdam UMC (approval code: NL55755.018.15).

### Mixed meal test

During the preoperative MMT, all participants orally ingested a standardized mixed meal (Nutridrink) after a fasting period of at least 9 hours. Blood sample collection, performed as described in the published study design,^25^ comprised seven time points.

### Fructose plasma measurements

On the day of the bariatric surgery, fasting peripheral blood samples were obtained prior to the procedure. Fasting plasma fructose concentrations were measured by Gas Chromatography – Triple Quadrupole Mass Spectrometry. Fructose was analyzed as the per-acetate derivative, prepared as previously described for glucose.^26^ D2-fructose (Campro, the Netherlands) was added as internal standard and both the M (Fructose) and the M^+2^ (D2-Frcutose) signal were monitored for accurate quantification by a 7890A GC, equipped with a 7693 auto sampler coupled to a 7000 Triple Quadrupole Mass Spectrometer (Agilent Technologies, U.S.A.).

### Ethanol plasma measurements

Endogenous ethanol concentrations were measured in plasma samples obtained during the MMT in the fasting state (t=0 min) and 120 minutes postprandially, as previously described.^24^ Ethanol measurements were performed in a subgroup of 34 individuals. Briefly, plasma samples were deproteinized and subsequently neutralized following centrifugation. Ethanol was initially assayed using an enzyme-based kit (DiaSys, Germany) and validated using high-performance liquid chromatography (HPLC) 4000 series (Jasco, Benelux). Ethanol was separated and next detected via refractive index detection. ChromNav chromatography software version 2.0 (Jasco, Benelux) was used for calculation of absolute ethanol concentrations. The difference between the fasting and postprandial ethanol concentration was used as a measure for postprandial ethanol increase.

### Tissue biopsies and histology

Intraoperative tissue biopsies were obtained from the liver and jejunum, as previously outlined.^25^ All tissue biopsies were either snap-frozen in liquid nitrogen and stored at -80°C until further processing, or formalin-fixed, paraffin-embedded for further histological analysis. Hematoxylin-eosin and Sirius red staining was performed on the paraffin-embedded biopsy sections. These histological sections were scored independently by two pathologists in accordance with the Steatosis, Activity, and Fibrosis score.^27^ Based on this scoring system, MASLD was characterized by isolated steatosis in more than 5% of hepatocytes or steatosis with mild inflammation, but without ballooning. MASH was characterized by steatosis in more than 5% of hepatocytes accompanied by both lobular inflammation and ballooning.

### RNA isolation and sequencing

Total RNA was extracted from the intraoperatively obtained liver and jejunum biopsies, homogenized, and purified. RNA quality was assessed on a BioAnalyzer instrument (Agilent Technologies, U.S.A.), and quantification was performed on Nanodrop (Thermo Fisher Scientific, U.S.A.). Due to RNA degradation, rRNA depletion was used for RNA-seq library preparation. Sequencing was performed at Novogene with 150-bp paired-end reads per sample. The average number of reads per sample was 42 ± 15 million. Obtained reads were quality-trimmed and pseudo-aligned to the human transcriptome. Full details are provided in a formerly published protocol.^24^

### Bulk RNA-seq analysis

Raw transcriptomic data from liver tissue was aggregated to gene-level counts using appropriate gene annotations. Differential gene expression analysis was performed using the DESeq2 package (v 1.50.2) in R, with fasting plasma fructose concentration included as a Z-score normalized continuous predictor. Batch effects were accounted for by including the batch variable as a covariate in the model design. No additional biological variables were included as covariates, to avoid obscuring associations with the primary predictor. Genes with fewer than 10 counts in at least two samples were excluded to improve data quality. Multiple testing correction was applied using the Benjamini-Hochberg false discovery rate (FDR) method. Genes with FDR adjusted *p*-values <0.05 were considered significantly differentially expressed. Functional enrichment of these genes was assessed using Gene Ontology (GO) enrichment analysis via the enrichGO() function and Kyoto Encyclopedia of Genes and Genomes (KEGG) pathway analysis via gseKEGG(), both conducted using the clusterProfiler package in R.

Following differential gene expression analysis, an expanded dataset comprising 289 BARIA individuals was used to evaluate hepatic expression of selected differentially expressed genes (DEGs) in relation to MASLD status. Raw liver gene counts were normalized using DESeq2 and transformed using a variance stabilizing transformation (VST) to reduce variability, particularly in low-count genes. VST-normalized expression levels were compared between MASLD and non-MASLD groups. For genes exhibiting significantly different expression between groups, binomial logistic regression models were applied to assess associations with MASLD (binary outcome), adjusting for age and sex as potential confounders. No additional variables were included to avoid overcorrection and loss of meaningful associations in this exploratory analysis.

To assess tissue-specific expression patterns of fructolysis genes within the study population, gene-level counts from both liver and jejunum were normalized using DESeq2, followed by VST. Genes of interest were extracted from the VST-normalized dataset for subsequent analysis and visualization.

### Single-nuclei RNA-seq analysis

To confirm our bulk RNA-seq findings in an independent cohort, we used publicly available single-nuclei RNA-seq data from Kim et al. (2025), containing 18 livers from healthy (normal), MASLD and MASH participants.^28^ We have downloaded the processed data (gene counts, gene and cell identifiers and metadata) from the original publication, and processed the data in Seurat, following the paper description, with minor modifications. Cell type allocation was performed using the same genes as the original paper. After subsetting only the normal and MASLD samples, we have conducted differential expression analysis with Seurat differential analysis functions, for the DEGs identified in the bulk RNA-seq that showed an association with MASLD. The findings were visualized as a volcano plot. Packages Seurat (v 5.3.0) and ggplot2 (v 4.0.0) were used for the analysis and visualization.

### Isolation of fecal total genomic DNA and microbiota profiling

Extraction of total fecal genomic DNA was performed using a modified IHMS extraction protocol^29^ involving homogenization and purification steps. Shotgun metagenomic sequencing libraries were prepared using a PCR-free method and sequenced with 150-bp paired-end reads per sample. Detailed description of the fecal total genomic DNA isolation is previously described.^24^ Adapter removal and quality trimming of reads were performed using fastp (v 0.23.4).^30^ Read quality was assessed using FastQC (v 0.12.1). Human reads were removed by mapping to the GRCh38.p14 human reference genome using Bowtie2 (v 2.2.5).^31^ The remaining non-human reads were profiled for microbial composition using the HUMAnN(v 3.9),^32^ MetaPhlan (v 4.1.1)^33^ pipelines.

### Fecal microbiota analysis

The phyloseq package (v 1.54.0) in R was used for preprocessing the microbiota data, including taxonomic agglomeration to the genus and species level. Fecal microbiota alpha diversity was quantified at the species level using the Shannon diversity index and its association with continuous fasting plasma fructose levels was assessed using Spearman rank correlation. Fecal microbiome beta-diversity at the species level was assessed using Bray–Curtis dissimilarity and associations with continuous fasting plasma fructose levels were tested with PERMANOVA (1000 permutations) using the vegan package (v 2.7-2). Subsequently, gut microbial species were filtered based on ≥20% prevalence across samples to minimize the risk of spurious associations in downstream analyses. Associations between fasting plasma fructose concentrations and the relative abundances of these microbial species were assessed using Spearman’s rank correlation coefficients with FDR correction. Statistical significance was defined as an FDR-adjusted *p* value < 0.05.

### *In vitro* bacterial culture experiments

*Streptococcus parasanguinis* DSM6778 and *Gordonibacter pamelaeae* DSM19378 were obtained from the Leibniz Institute DSMZ (DSMZ GmbH, Germany). Both strains were cultivated at 37°C under anaerobic conditions (5% H_2_, 10% CO_2_, 85% N_2_) in YCFA media at pH 6.5 (10g/l casein hydrolysate, 10g/l yeast extract, 4g/l NaHCO_3_, 2.7g/l sodium acetate, 4.5g/l K_2_HPO_4_, 0.7g/l KH_2_PO_4_, 0.9g/l (NH_4_)_2_SO_4_, 0.9g/l NaCl, 0.09g/l MgSO_4_, 0.09g/l CaCl_2_, 0.5g/l L-cysteine, 1µg/l biotin, 1µg/l cobalamin, 3µg/l para-aminobenzoic acid, 5µg/l folic acid, 15µg/l pyridoxamine) with the addition of 10mM fructose as a carbon source for *S. parasanguinis* DSM6778 or the addition of 1% of L-arginine for *G. pamelaeae* DSM19378. The strains were grown in triplicate on single culture or mixed culture at a 1:1 ratio for 48h in a 96-well plate setting using YCFA media with the addition of 1% L-arginine for all conditions and varying fructose concentrations (no addition, 10mM or 20mM). Inoculation ratio was 5% for both tested strains from an active culture. Growth was monitored by measuring optical density at 600nm (OD_600_) during incubation at 37°C for 48h using a Biotek Epoch 2 microplate reader (BioTek, USA).

Samples were collected at 0 and 48 hours of culture across all tested conditions and were centrifuged at 18.000 rpm for 5 minutes to precipitate microbial cells. The supernatant was analyzed for metabolites using HPLC using a SH1821 (SUGAR) column (Shodex, Japan). The mobile phase was sulfuric acid 0.05 N with a flow rate of 1mL/min. The column was kept at a temperature of 50°C for RI and UV (210nm) measurements. Appropriate standards were used for the identification and quantification of metabolites.

### Statistical analysis

Although this was an exploratory study in an ongoing cohort study, a power calculation was performed. Sample size calculation was based on a previous publication reporting a mean plasma fructose level of 46±25.22 μM in samples from subjects being evaluated for diabetes control.^34^ Using a two-sample t-test for two independent groups (with a two-sided significance level of 0.05 and 80% power), we estimated that a sample size of approximately 38 subjects would be required to detect a 50% difference in plasma fructose concentrations between non-MASLD and MASLD subjects. As unequal group sizes are expected, a total of approximately 48 subjects would therefore be required to maintain adequate statistical power. Descriptive statistics for continuous variables following a normal distribution were expressed as mean ± Standerd Deviation (SD) and non-normally distributed variables as median Inter Quartile Range [IQR]. Normality was assessed using the Shapiro-Wilk test. Group-comparisons for normally distributed variables were performed using unpaired t-tests, and non-normally distributed variables were assessed using the Wilcoxon rank-sum test. The table 1 was created using the tableone package (v 0.13.2). Correlation analyses were conducted using Spearman’s rank correlation. Statistical significance was set at a two-tailed p-value <0.05. All statistical analysis were performed using R (version 4.5.2). The underlying code for this study is not publicly available but may be made available to qualified researchers on reasonable request from the corresponding author.

**Table 1.**
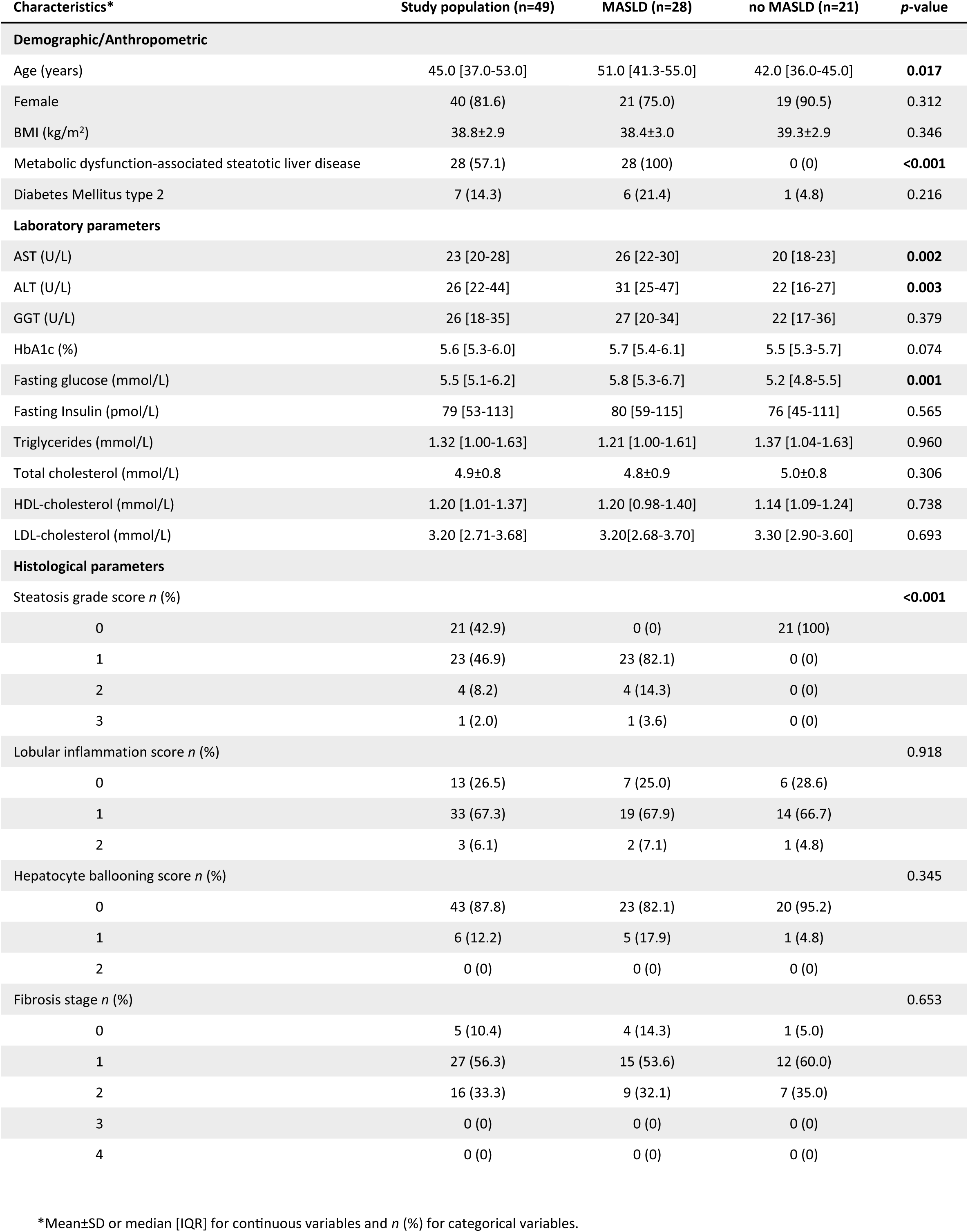
Baseline characteristics. *Mean±SD or median [IQR] for continuous variables and *n* (%) for categorical variables.

## Results

### Patient baseline characteristics

This study included a subset of 49 morbidly obese subjects enrolled in the prospective BARIA cohort, with data collected prior to and during bariatric surgery. Morbid obesity was defined as a body mass index (BMI) ≥40 kg/m2 or a BMI ≥35 kg/m2 with obesity-related comorbidity. The baseline characteristics (Table 1) were stratified by MASLD status. Overall, the majority of this study population, 81.6%, was female and 57.1% of the subjects had MASLD. The median [IQR] age was 45.0 [37.0-53.0] years, and the mean BMI±SD before surgery was 38.8±2.9 kg/m^2^. Subjects with MASLD were significantly older than those without MASLD.

### MASLD is associated with higher fasting plasma fructose levels

In contrast to the well-established link between high dietary fructose intake and MASLD, the association between fasting plasma fructose levels and MASLD remains poorly characterized. To investigate this, we compared fasting peripheral plasma fructose concentrations between subjects with and without MASLD. Subjects with MASLD showed significantly higher fasting fructose levels with a median [IQR] of 20.32 [17.45-23.82] µM, compared to 16.57 [15.77-20.33] µM in non-MASLD subjects (Figure 1a). Differential gene expression analysis of liver bulk RNA-seq data did not demonstrate upregulation of polyol pathway genes in association with increased fasting plasma fructose levels, with *SORD* expression even showing a significant downregulation (Figure 1b). Among lipogenic genes, *MLXIPL* – encoding transcription factor ChREBP – was marginally upregulated and *FASN*, the main regulator of hepatic FA synthesis, was significantly upregulated (Figure 1c). This is consistent with the well-established increase in DNL induced by high-fructose intake.^35–41^ Furthermore, there was no significant correlation between fasting fructose and glucose concentrations. Collectively, these findings suggest that fasting plasma fructose levels do not reflect endogenous production by glucose conversion through the polyol pathway, but are rather indicative of (prolonged) high dietary intake.

**Figure 1.**
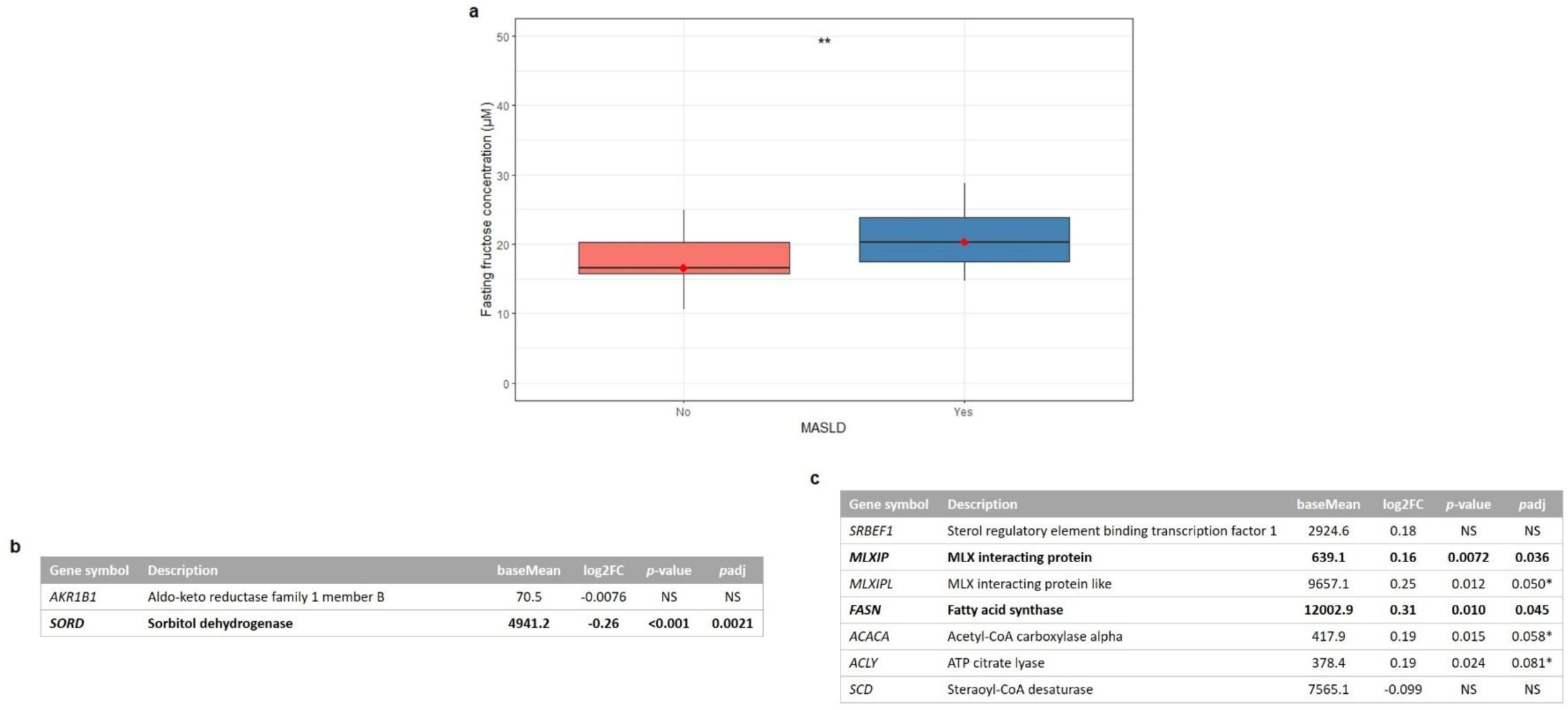
Fasting plasma fructose levels are higher in MASLD subjects. **(a)** Fasting fructose concentrations stratified by MASLD status (no MASLD, *n* = 21; MASLD, *n* =28) \*\**p*<0.01. **(b)** Hepatic DESeq2 analysis results of polyol pathway genes in relation to fasting plasma fructose concentrations. Expression of *SORD* was significantly downregulated in relation to increased fasting fructose levels. (**c**) Hepatic DESeq2 analysis results of lipogenic genes in relation to fasting plasma fructose concentrations. The results show upregulation of *FASN* and ChREBP encoding genes. **\***no significant *p*-value after correction for multiple testing. Log2FC, log2 fold change; *p*adj: adjusted *p*-value.

### Fasting plasma fructose concentrations positively correlate with postprandial ethanol excursions

Recent literature proposes that fructose toxicity arises in part from its conversion into ethanol by gut microbial fermentation.^23^ Hence, we investigated the relationship between fasting plasma fructose levels and the postprandial increase in plasma ethanol concentrations. Plasma ethanol levels were measured in MMT samples obtained in a fasting state and at 120 minutes postprandially. A significant positive linear correlation between fasting plasma fructose levels and postprandial plasma ethanol excursions was identified (Figure 2a). Importantly, this correlation was independent of reported exogenous alcohol consumption (Supplementary Figure 1). This indicates that increased fasting plasma fructose concentrations, as a potential proxy of high dietary fructose exposure, may reflect enhanced microbial ethanol production capacity due to intestinal microbiota alterations and/or altered ethanol catabolism. The postprandial increase in plasma ethanol was significantly higher in MASLD subjects (median [IQR]: 35.83 [29.11–56.30] µM), than in non-MASLD subjects, (14.73 [-1.26–20.78] µM) (Figure 2b).

**Figure 2.**
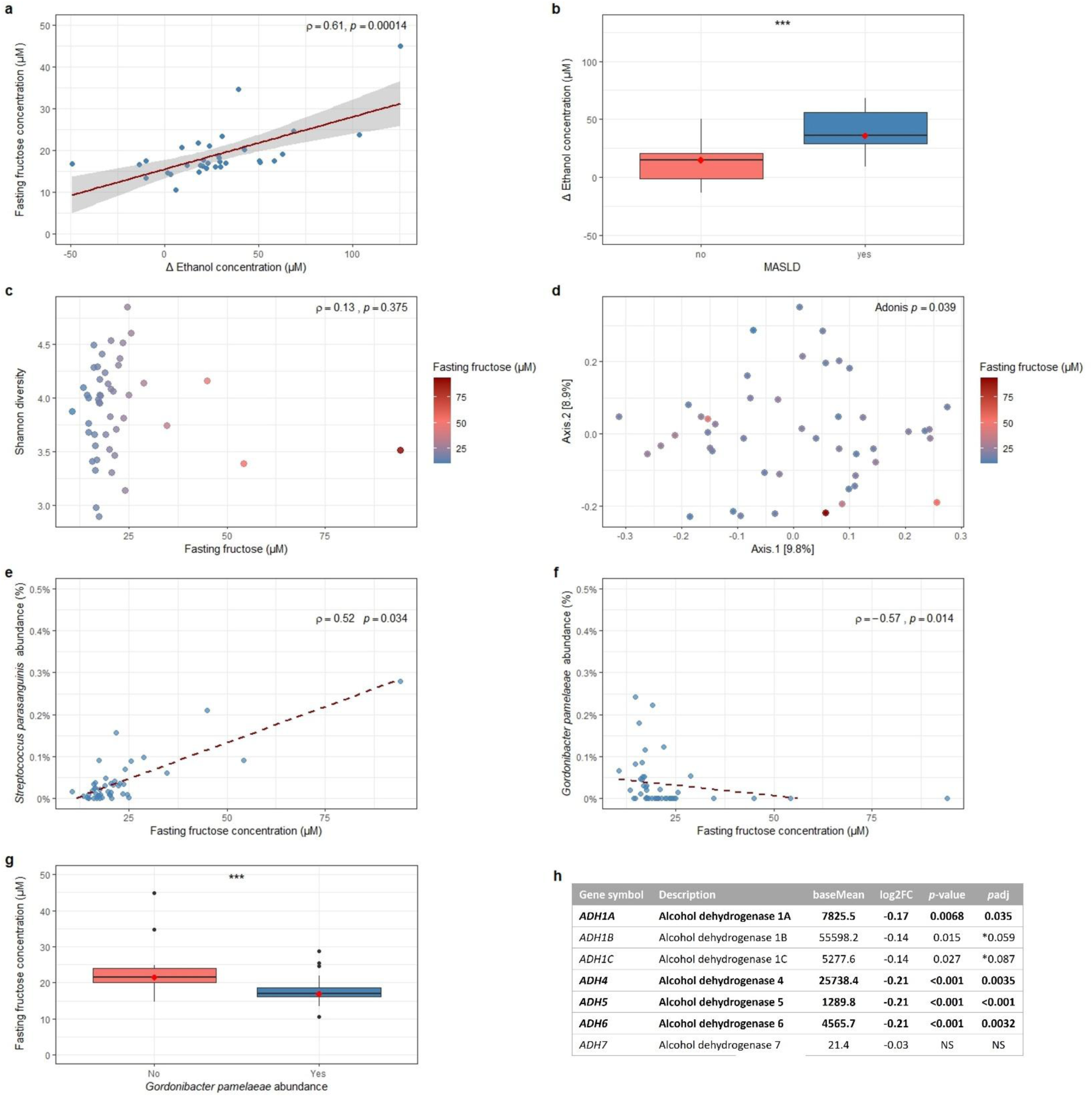
Fasting plasma fructose concentrations are positively correlated with postprandial plasma ethanol excursions. **(a**) Correlation between fasting plasma fructose concentrations and postprandial ethanol excursions. (**b**) Δ Ethanol concentrations stratified by MASLD status (no MASLD, *n* = 21; MASLD, *n* = 28) \*\*\**p*<0.001. In (**a**) and (**b**), Δ Ethanol concentration is the difference between the concentrations measured 120 minutes postprandial and in the fasting state during a mixed meal test. (**c**) Alpha diversity was quantified using Shannon index. (**d**) Beta diversity was assessed using Bray-Curtis dissimilarity and PERMANOVA (1000 permutations). (**e**,**f**) Correlation between fasting plasma fructose levels and the relative abundance of *Streptococcus parasanguinis* (**e**) and *Gordonibacter pamelaeae* (**f**). Spearman’s rho correlations between fasting fructose levels and gut microbial species were adjusted for multiple testing using FDR correction (**e**,**f**). (**g**) Fasting fructose concentrations in subjects with and without presence of *Gordonibacter* pamelaeae in the fecal microbiota (yes, *n* = 26; no, *n* = 21) \*\*\**p*<0.001. (**h**) Hepatic DESeq2 analysis results of ADH encoding genes in relation to fasting plasma fructose concentrations. *ADH1A*, *ADH4*, *ADH5*, *ADH6* were significantly downregulated in relation to increased fasting fructose levels. **\***no significant *p*-value after correction for multiple testing. Log2FC, log2 fold change; *p*adj: adjusted *p*-value.

### Correlations with fecal microbiota composition and ethanol catabolism at the transcriptomic level

To elucidate the mechanism underlying the link between fasting plasma fructose levels and postprandial plasma ethanol increase, we first assessed the overall gut microbiota composition in relation to fasting fructose. Fecal microbiota alpha-diversity did not show a significant association with fasting plasma fructose concentrations (Figure 2c). In contrast, fecal microbial beta-diversity was significantly associated with fasting fructose levels (Figure 2d), explaining 3.1% of variance, indicating distinct fecal microbiota compositions between subjects with high and low fasting fructose concentrations. Next, we assessed correlations between fasting fructose levels and the relative abundance of overall fecal microbiota species. Among all species analyzed, fasting fructose positively correlated with *Streptococcus parasanguinis* (Figure 2e), and negatively correlated with *Gordonibacter pamelaeae* (Figure 2f). This negative correlation was primarily influenced by higher fasting fructose concentrations among subjects lacking detectable *G. pamelaeae* abundance (Figure 2g). Correlations with other microbial species, as well as functional pathways were also assessed but did not reach statistical significance.

In addition to these correlations, bulk RNA-seq data showed a downregulation of multiple alcohol dehydrogenase (ADH) encoding genes in relation to increased fasting plasma fructose levels (Figure 2h). This indicates that, besides potential enhanced endogenous ethanol production, alterations in ethanol catabolism coincide with the greater postprandial ethanol excursions observed in subjects with higher fasting plasma fructose levels.

### *In vitro* growth and metabolite profiles of *S. parasanguinis* and *G. pamelaeae* in response to fructose

To investigate the correlation between fasting fructose and the two associated species, we performed *in vitro* experiments with *S. parasanguinis* and *G. pamelaeae* grown individually and in co-culture across two fructose concentrations. *S. parasanguinis* exhibited dose-dependent growth stimulation by fructose (Figure 3a), supporting its role as a fructose fermenter. *G. pamelaeae*, by contrast, showed no growth response to fructose (Figure 3b), consistent with its negative correlation with fasting fructose in our cohort. In co-culture, a dose-dependent fructose response was retained, though with an accelerated decline phase compared to both monocultures (Supplementary Figure 2).

**Figure 3.**
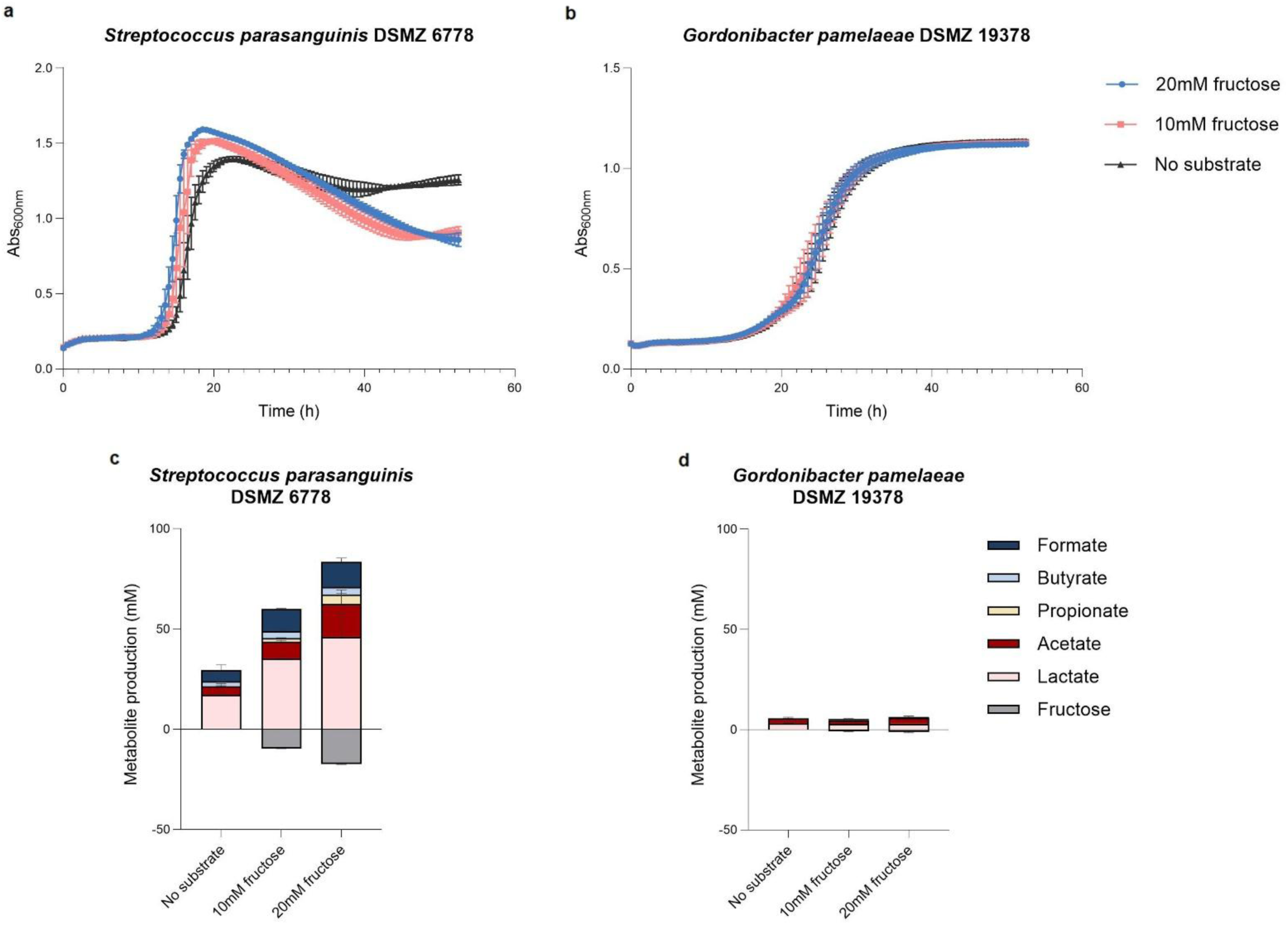
Fructose-dependent growth dynamics and fermentation profiles of *S. parasanguinis* and *G. pamelaeae in vitro*. (**a-b**) Growth curves (Abs₆₀₀ₙₘ) of *S. parasanguinis* (**a**) and *G. pamelaeae* (**b**) at 0, 10, and 20 mM fructose. (**c-d**) Fermentation end-products of *S. parasanguinis* (**c**) and *G. pamelaeae* (**d**) across fructose concentrations.

Metabolite profiling revealed distinct, species-dependent fermentation profiles (Figure 3c,d). In monoculture, *S. parasanguinis* produced lactate as its primary fermentation end-product in a fructose dose-dependent manner (Figure 3c), with acetate and formate as the most abundant secondary products, consistent with its classification as a lactic acid bacterium. In contrast, *G. pamelaeae* generated only low metabolite levels overall (Figure 3d), reflecting its limited fructose-fermenting capacity. Ethanol was not detected under any condition.

### Hepatic and jejunal expression of fructolysis genes in a morbidly obese population

Both the liver and jejunum are tissues involved in fructose catabolism and exhibit high expression levels of fructolysis-related genes. Nevertheless, consensus expression data are usually derived from non-diseased tissues, which might not accurately reflect the tissue-specific expression profiles in a morbidly obese population. To obtain further insights into the involvement of the liver and jejunum in fructose catabolism among morbidly obese subjects, the expression of the genes encoding the main enzymes involved in fructolysis was compared across these tissues. VST-normalized expression levels of *KHK*, *ALDOB*, and *TKFC* were analyzed (Figure 4). In this population, *KHK* expression was highest in liver, whereas *TKFC* expression was highest in jejunum, consistent with consensus expression patterns reported by the Human Protein Atlas.^42,43^ *ALDOB* showed higher expression in jejunum than in liver contrasting Human Protein Atlas data showing greater hepatic expression.^44^ Induction of *Aldob* expression in jejunum following fructose feeding has been observed in mice,^45^ suggesting that jejunal *ALDOB* expression could be upregulated in a morbidly obese population as a metabolic adaptation to increased fructose exposure.

**Figure 4.**
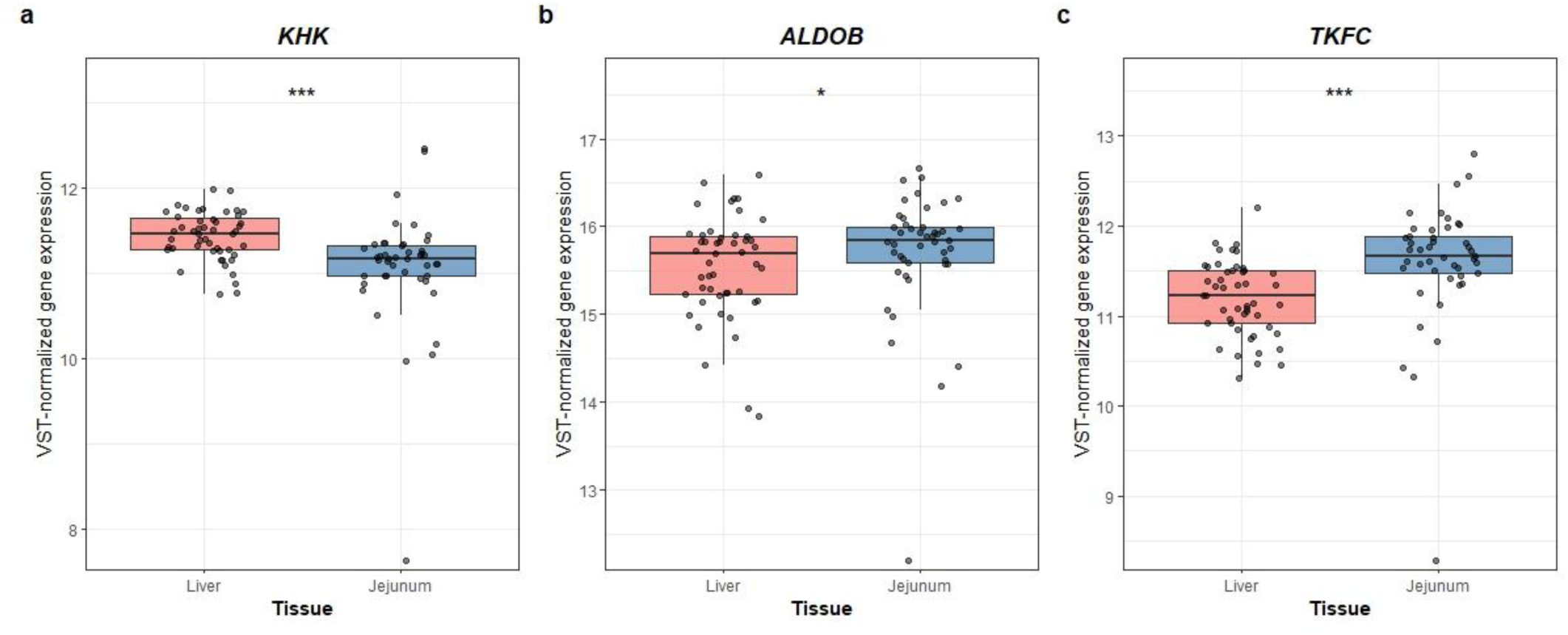
Hepatic and jejunal expression levels of fructolysis genes in a morbidly obese population. (**a-c**), VST-normalized gene counts of fructolysis genes stratified by tissue type (liver and jejunum), including *KHK* (**a**), *ALDOB* (**b**), *TKFC* (**c**). \**p*<0.05, *** *p*<0.001. VST, variance stabilizing transformation.

### Increased fasting plasma fructose levels are associated with extensive hepatic transcriptional changes

To gain a deeper understanding of the pathways and specific genes underlying the association between fasting plasma fructose levels and MASLD, transcriptional changes related to increased fasting fructose concentrations were analyzed using DESeq2. As the liver is the primary target of fructose toxicity, the analysis focused on hepatic transcriptomics. 3969 significant DEGs were identified in liver tissue, of which 1522 were found to be upregulated and 2447 downregulated (Figure 5a). When applying a minimum absolute Log2FoldChange ≥1, this resulted in one upregulated and 19 downregulated DEGs. No distinct clustering of samples or gene expression patterns were observed among these DEGs in relation to fasting fructose levels (Supplementary Figure 3). Importantly, genes exhibiting smaller but statistically significant changes may still be crucial in driving meaningful transcriptional effects.

**Figure 5.**
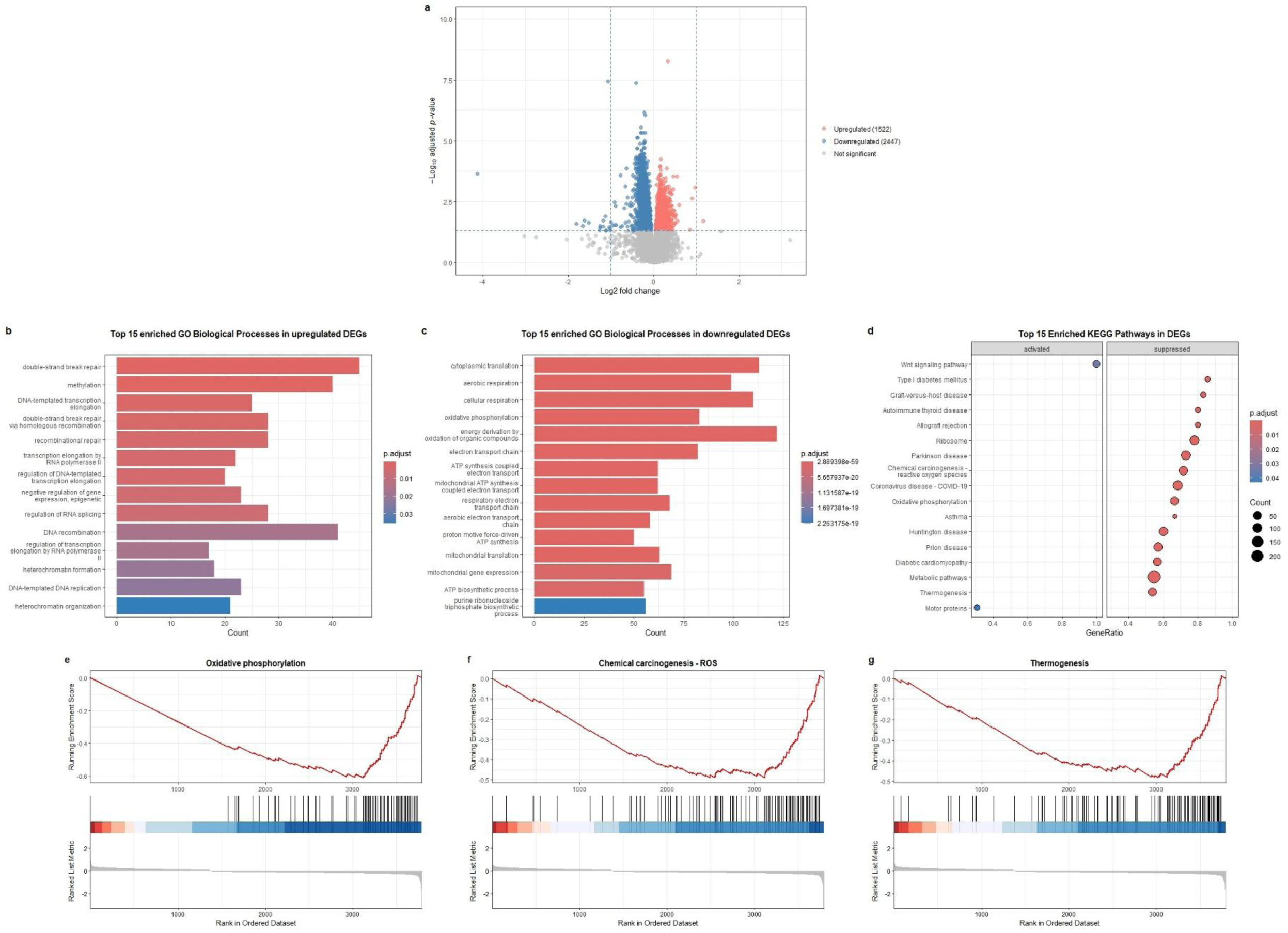
RNA-seq differential gene expression and enrichment analysis. (**a**) Volcano plot illustrating the distribution of DEGs in association with increased fasting fructose levels. (**b**) Top 15 enriched GO biological processes in the upregulated DEGs. (**c**) Top 15 enriched GO biological processes in the downregulated DEGs. (**d**) Top 15 enriched KEGG pathways in the DEGs. (**e**) Enrichment plot of the oxidative phosphorylation KEGG pathway (NES= -3.20, *p*<0.001). (**f**) Enrichment plot of the chemical carcinogenesis – reactive oxygen species KEGG pathway (NES= -2.69, *p*<0.001). (**g**) Enrichment plot of the thermogenesis KEGG pathway (NES= - 2.65, *p*<0.001). DEGs, differentially expressed genes; GO, gene ontology.

### Functional enrichment analysis associates increased fasting plasma fructose levels with impaired mitochondrial energy metabolism and antioxidant defense

GO Biological Process enrichment analysis on all DEGs demonstrated that upregulated genes were significantly enriched in processes related to DNA repair, transcriptional regulation and epigenetic modification (Figure 5b). Downregulated genes were predominantly enriched in processes related to mitochondrial function and energy metabolism (Figure 5c). Among the top 15 enriched KEGG pathways (Figure 5d), suppression of the oxidative phosphorylation (OXPHOS) pathway was identified (Figure 5e), consistent with GO enrichment results showing downregulation of processes involved in mitochondrial energy metabolism. Examination of the KEGG OXPHOS pathway map (hsa00190) showed widespread downregulation of genes across all major electron transport chain (ETC) complexes (Supplementary Figure 4). All OXPHOS pathway DEGs were extracted and their DESeq2 results summarized, confirming broad impairment of mitochondrial energy metabolism (Supplementary Table 1).

Given the effects on mitochondrial energy metabolism, the opposing yet interconnected roles of β-oxidation and lipogenesis, and the role of fructose as a hepatic lipogenic substrate, downstream β-oxidation genes were also examined (Supplementary Table 2). The majority of mitochondrial β-oxidation genes were significantly downregulated, further highlighting the previously observed impaired mitochondrial energy metabolism. Notably, *ACADM* expression was significantly reduced, *ACADVL* was upregulated, suggesting a shift suggesting a shift towards the oxidation of long-chain fatty acids (LCFA).

Moreover, a suppression of the chemical carcinogenesis – reactive oxygen species (ROS) KEGG pathway was found (Figure 5d,f), suggesting alterations in the cellular oxidative stress response. The KEGG pathway map (hsa05208) revealed downregulation of key genes involved in antioxidant defense and cellular detoxification (Supplementary Figure 5). Detailed analysis of liver-expressed antioxidant genes showed significantly reduced expression of *SOD1*, *CAT* and nearly all components of the glutathione pathway, thioredoxin system, peroxiredoxin system and AKR family (Supplementary Table 3).

Downregulation of the thermogenesis pathway was also identified (Figure 5d,g). Although uncoupling protein 1 (UCP1) – the mitochondrial protein central to thermogenesis – is not expressed in liver, downregulation of this pathway could reflect a broader suppression of mitochondrial activity and energy metabolism. Pathway map analysis (hsa04714) illustrated the previously observed downregulation of OXPHOS genes, alongside reduced expression of genes encoding protein kinase A (PKA) and its downstream target *CREB1* (Supplementary Figure 6). *CREB1*, in turn, can induce expression of *PPARGC1A*, encoding the master regulator of mitochondrial biogenesis, whose expression was similarly downregulated. Targeted analysis confirmed significant downregulation of *PPARGC1A, CREB1* and nearly all genes encoding PKA subunits (Supplementary Table 4), indicating coordinated suppression of this metabolic signaling pathway that affects mitochondrial function. Together, these functional enrichment analyses results demonstrate that higher fasting plasma fructose levels are associated with mitochondrial dysfunction, encompassing impaired mitochondrial energy metabolism and antioxidant defense, in a morbidly obese population.

### Gene expression changes in mitochondrial energy metabolism and antioxidant defense link elevated fasting fructose to increased MASLD risk

Expression of genes involved in mitochondrial energy metabolism and antioxidant defense were associated with fasting plasma fructose levels, which in turn were linked to MASLD histology scores. Therefore, we investigated whether these DEGs were differentially expressed by MASLD status. Gene expression was assessed in an expanded subset of the population (Supplementary Table 5), stratified by histology-based MASLD status. The analysis included all identified DEGs related to OXPHOS, β-oxidation, antioxidant defense and the regulatory genes encoding PKA, *CREB1,* and *PPARGC1A*.

Among downregulated OXPHOS pathway genes, expression levels of *NDUFA5* and *NDUFB5*, *UQCRH* and *COX7A2* were significantly lower in MASLD subjects compared to non-MASLD subjects (Figure 6a-d). In logistic regression analysis, lower expression of each of these genes was independently associated with increased MASLD risk, after adjusting for the confounders age and sex (Supplementary Figure 7). Expression of β-oxidation genes *ACADM* and *ACADVL* also differed significantly between groups (Figure 6e,f). *ACADM* expression was lower in MASLD subjects, whereas *ACADVL* was higher, consistent with their expression changes related to elevated fasting fructose. Reduced *ACADM* expression was marginally associated with increased MASLD risk, while increased *ACADVL* expression was significantly associated with a higher MASLD risk (Supplementary Figure 7). Of the differentially expressed antioxidant defense genes, *SOD1*, *PRDX6* and *AKR1C4* expression levels were all significantly lower in MASLD subjects (Figure 6g-i). Lower expression of SOD1 and PRDX6 were each significantly associated with increased odds of MASLD, whereas AKR1C4 expression was not associated with MASLD risk (Supplementary Figure 7). Among the identified regulatory DEGs, the expression of *PRKAR1A*, which encodes the most highly expressed PKA regulatory subunit in liver, was significantly lower in MASLD subjects (Figure 7a), as were *CREB1* and *PPARGC1A* expression (Figure 7b,c). Lower expression of *PRKAR1A* and *PPARGC1A* were each strongly associated with increased MASLD risk (Supplementary Figure 7), whereas *CREB1* expression was not.

**Figure 6.**
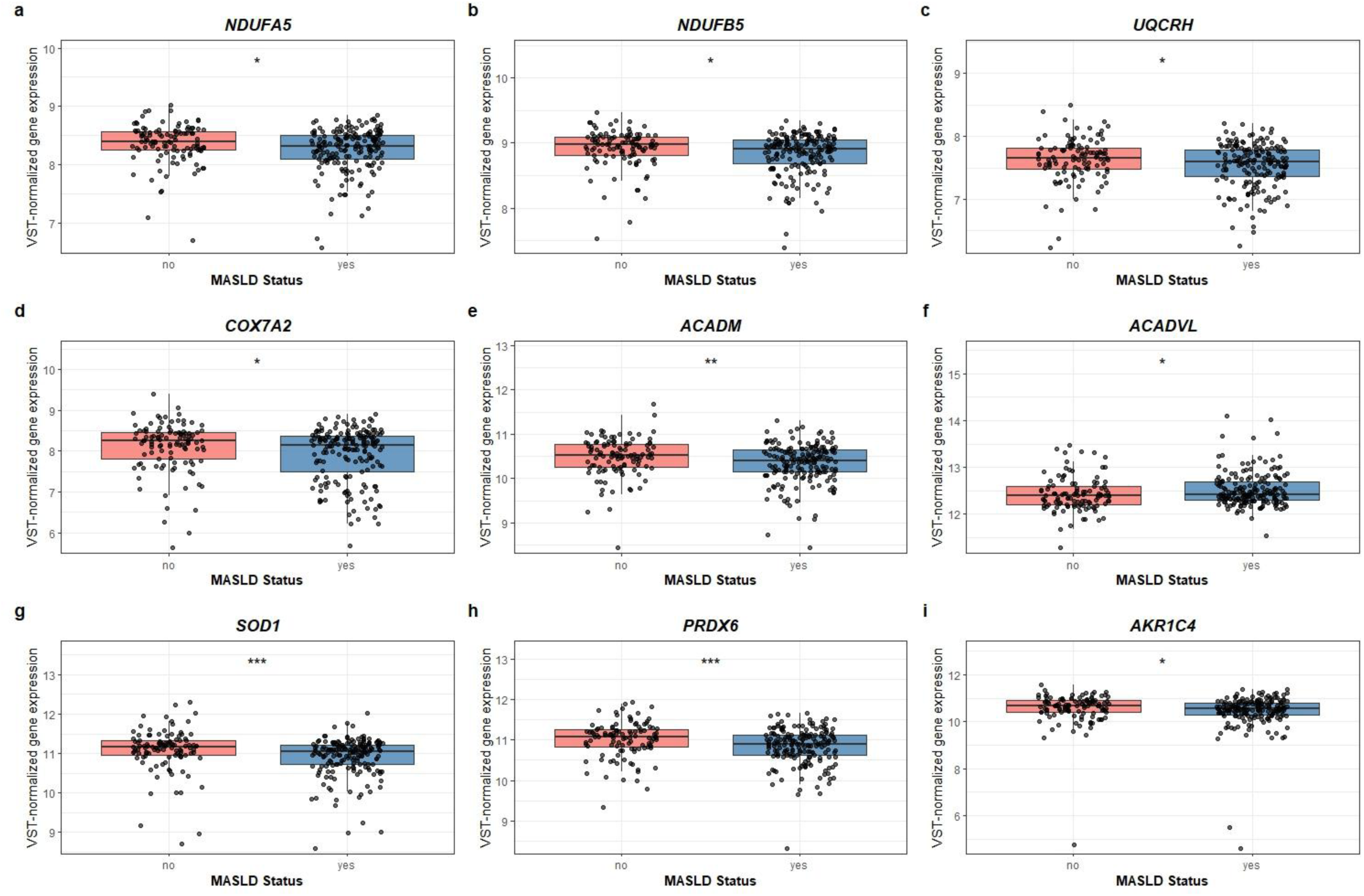
Altered hepatic expression of genes involved in oxidative phosphorylation, mitochondrial β-oxidation, and antioxidant defense in MASLD subjects. (**a**-**i**) VST-normalized hepatic expression of DEGs involved in oxidative phosphorylation, mitochondrial β-oxidation and antioxidant defense in an expanded subset of the cohort (*n* = 289), stratified by MASLD status (no MASLD, *n* = 109; MASLD, *n* = 180). DEGs exhibiting significantly different expression between MASLD and non-MASLD subjects are illustrated, including *NDUFA5*, *p*=0.010 (**a**), *NDUFB5*, *p*=0.024 (**b**), *UQCRH* (*p*=0.048) (**c**), *COX7A2*, *p*=0.017 (**d**), *ACADM*, *p*=0.0040 (**e**), *ACADVL*, *p*=0.019 (**f**) *SOD1*, *p*<0.001 (**g**), *PRDX6*, *p*<0.001 (**h**), *AKR1C4*, *p*=0.024 (**i**). \**p*<0.05, \*\**p*<0.01, *** *p*<0.001. VST, variance stabilizing transformation.

**Figure 7.**
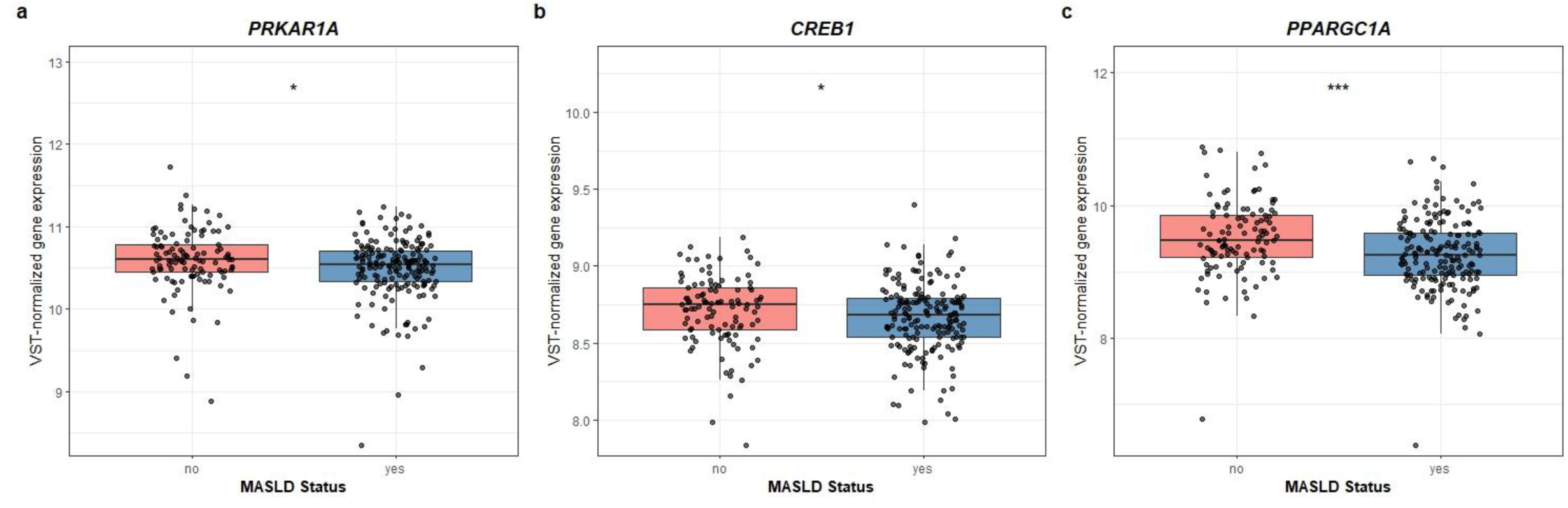
Lower hepatic expression of *PRKAR1A*, *CREB1* and *PPARGC1A* in subjects with MASLD. (**a**-**c**) VST-normalized hepatic expression of all regulatory genes encoding PKA, *CREB1* and *PPARGC1A* was compared between all subjects of the study population, stratified by MASLD status. Genes exhibiting significantly different expression between MASLD and non-MASLD subjects are demonstrated, including *PRKAR1A*, *p*=0.016 (**a**), *CREB1,* p=0.044 (**b**), PPARGC1A, *p*<0.001 (**c**). \**p*<0.05, \*\*\**p*<0.001.

Collectively, these results suggest that downregulation of multiple OXPHOS and antioxidant defense genes, as well as dysregulation of β-oxidation genes, contribute to increased MASLD risk. The directionality of these associations was consistent with fasting fructose-associated gene expression changes. This indicates that impaired mitochondrial energy metabolism and antioxidant defense, potentially influenced by downregulation of PKA and *PPARGC1A* signaling, may represent a mechanistic link between increased fasting plasma fructose levels and MASLD pathogenesis.

### Single-nuclei RNA-seq analysis highlights cell-specific downregulation of *PPARGC1A* in MASLD

To investigate the gene expression patterns of the DEGs associated with MASLD across distinct cell types, we re-analyzed single-nuclei RNA-seq data from an independent cohort.^28^ This analysis included the subset of DEGs that were associated with MASLD in our bulk RNA-seq findings, in addition to differentially expressed lipogenic genes, comparing MASLD subjects to controls. We found significant downregulation of *PPARGC1A* in hepatocytes, endothelial cells, and myeloid cells (Figure 8), whereas *FASN* was markedly upregulated in these same cell types, as well as in B cells, T cells, and hepatic stellate cells (HSCs). Apart from the well-established role of FASN as a lipogenic gene in MASLD, these results further highlight the potential contribution of PPARGC1A downregulation — associated with elevated fasting plasma fructose — to MASLD pathogenesis. Based on the affected cell types, suppression of *PPARGC1A* may be associated not only with disruptions in metabolic homeostasis, but also with disturbances in vascular homeostasis and immune responses.

**Figure 8.**
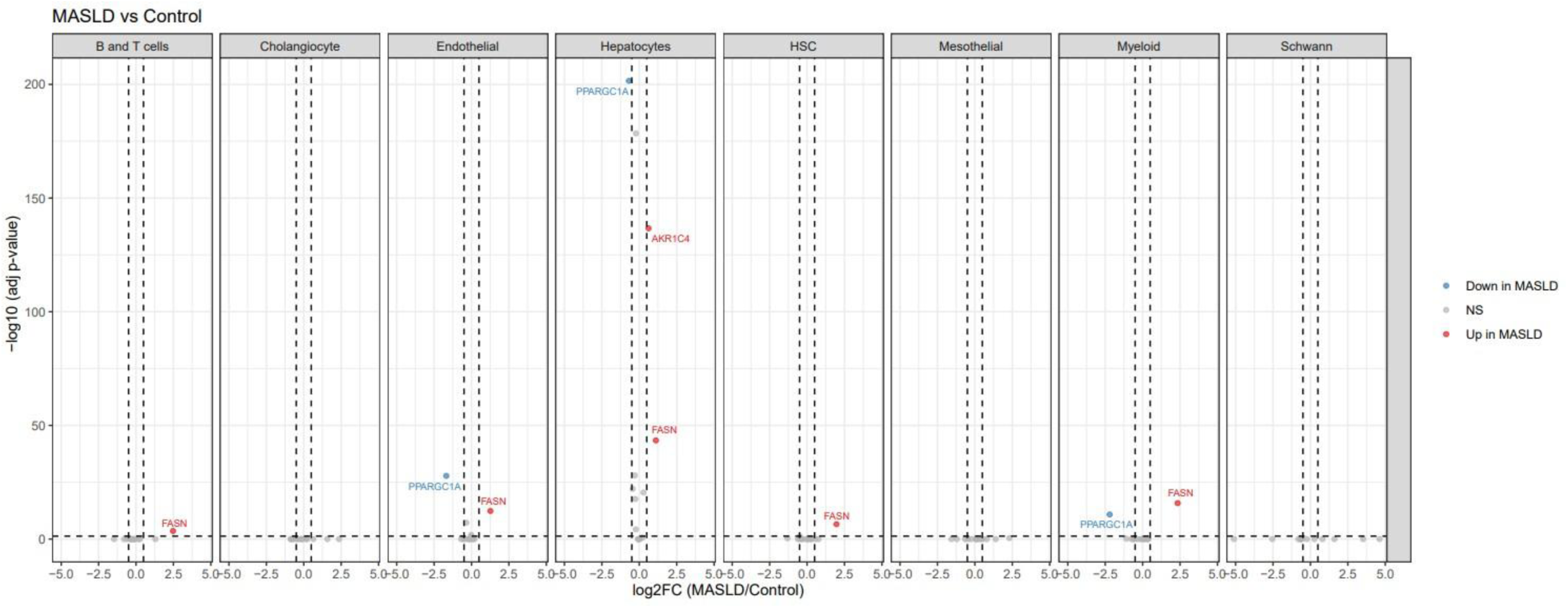
Single-nuclei RNA-seq data highlights *PPARGC1A* downregulation in various hepatic cell-types in MASLD subjects. Differential expression analysis in single-nuclei RNA-seq data, comparing MASLD versus control samples. DEGs identified in bulk RNA-seq analysis, that were associated with MASLD, were included in the analysis. Log2FC, log2 fold change; NS, not significant.

## Discussion

High dietary fructose drives MASLD development, but the significance of fasting plasma fructose levels in relation to MASLD remains to be elucidated. Using plasma metabolites, fecal shotgun metagenomic data, and hepatic bulk- and single-nuclei RNA-seq data, we explored associations with fasting plasma fructose and their link to MASLD. We found significantly higher fasting plasma fructose concentrations in individuals with MASLD compared to those without. Fasting plasma fructose levels positively correlated with postprandial plasma ethanol excursions, which may result from microbial fructose fermentation, suggesting shifts in gut microbial composition and/or changes in host ethanol catabolism. This was further supported by associations between fasting fructose and gut microbiota composition, including a significant effect on beta-diversity and correlations with the abundances of *S. parasanguinis* and *G. pamelaeae*, as well as downregulation of hepatic ADH-encoding genes with increased fasting fructose concentrations. Finally, transcriptomic analysis revealed that heightened fasting plasma fructose levels are associated with impaired hepatic mitochondrial energy metabolism and antioxidant defense.

One previous cohort study linked elevated fasting plasma fructose to increased MASLD risk, hypothesizing it reflects endogenous production.^46^ However, downregulation of *SORD* in our study suggests reduced conversion of sorbitol to fructose, challenging this hypothesis. Moreover, there was no correlation between fasting fructose and glucose in our study. This lack of correlation is not unexpected, given that fructose catabolism follows a distinct metabolic pathway and is largely independent of systemic glucose homeostasis.^47^ Taken together with the upregulation of lipogenic genes, our findings are more consistent with a hepatic effect associated with increased dietary fructose exposure.

The gut microbiota can endogenously produce clinically relevant amounts of ethanol through sugar fermentation, which is implicated in MASLD pathogenesis.^24,48,49^ The positive correlation between fasting plasma fructose and postprandial ethanol excursions implies that higher fasting fructose reflects a gut microbial composition more prone to ethanol production. Beta-diversity analysis confirmed distinct fecal microbiota compositions between subjects with low and high fasting fructose levels. Previous studies have shown that high-fructose intake alters microbial profiles and promotes aberrant gut microbiota composition^50^ and *in vitro* data confirm fructose as a suitable substrate for ethanol production.^51^ Together, this suggests that dietary fructose-driven modulation of the gut microbiota may explain the correlation between fasting plasma fructose and postprandial ethanol production, supporting a role for fructose as a substrate for endogenous ethanol production *in vivo*. Furthermore, high circulating (endogenously produced) plasma ethanol has previously been ascribed to TNFα-dependent impairment of ADH activity,^52^ consistent with our observed downregulation of ADH encoding genes in relation to high fasting plasma fructose, This may contribute to the positive correlation between fasting fructose and postprandial ethanol excursions.

*S. parasanguinis* was positively correlated with fasting plasma fructose levels, while *G. pamelaeae* was inversely correlated, yet neither species has been identified as an ethanol producer.^22^ Despite limited literature on their abundance in relation to fructose exposure, both species have been described in the context of MASLD. S*. parasanguinis* is a typical oral commensal and enrichment of oral microbiota has been observed in females with MASLD.^53^ Correspondingly, *S. parasanguinis* was among the top discriminatory species differentiating MASLD-cirrhosis from non-MASLD controls,^54^ and positively correlated with polyamines,^53^ whose excessive catabolism can drive oxidative stress, which is implicated in the development of MASLD.^55–57^ *G. pamelaeae* converts ellagic acid into urolithin C,^58^ a metabolite that protects against MASLD by maintaining intestinal barrier integrity and mitigating gut dysbiosis in a murine model.^59^ Accordingly, this species was negatively associated with MASLD and predicted healthy status in overweight individuals,^60^ further supporting a protective role. Our *in vitro* experiments confirm that *S. parasanguinis* ferments fructose, consistent with its positive correlation with fasting fructose, producing lactate and acetate as major fermentation end-products in a fructose dose-dependent manner. As expected, no ethanol was detected. Nevertheless, these fermentation products may potentiate the hepatotoxic effects of fructose by serving as substrates for hepatic de novo lipogenesis,^21^ or indirectly as cross-feeding substrates for bacteria capable of producing toxic metabolites such as ethanol. Collectively, increased *S. parasanguinis* and decreased *G. pamelaeae* abundance are associated with pathogenic effects linked to MASLD and with gut dysbiosis, which may reflect unrecognized enrichment of ethanol-producing strains within the broader microbial community.

Dietary fructose exerts metabolic effects beyond promoting hepatic lipogenesis. In rodent models, high-fructose diets primarily impaired mitochondrial function including β-oxidation,^61,62^ and OXPHOS was the most significantly downregulated pathway in mice fed a high-fructose, high-fat diet, compared to a high-fat diet alone.^63^ Consistently, our study revealed downregulation of many hepatic genes involved in β-oxidation and OXPHOS. Moreover, key antioxidant genes including *CAT*, *SOD1*, glutathione peroxidases (GPx), thioredoxins and periredoxins were downregulated. Animal studies similarly showed impaired hepatic antioxidant defense in high-fructose fed rats, demonstrated by reduced mRNA and protein expression of Cat, Sod1 and GPx,^64–66^ along with decreased mitochondrial Sod1 and Cat enzyme activities.^67,68^ In our study, the expression of multiple genes involved in these processes was linked to MASLD risk. This aligns with previous studies that identified mitochondrial dysfunction as a driver of MASLD pathogenesis and progression,^69–73^ with impaired OXPHOS, β-oxidation, and antioxidant defense playing key roles particularly in advanced disease stages.^69–73^ Mitochondrial dysfunction, reflected by reduced β-oxidation and carnitine palmitoyltransferase 1 (CPT1) activity, preceded MASLD development in a rat model^74^, also emphasizing the role of early mitochondrial impairment in disease onset. *ACADVL* upregulation and its association with MASLD in the present study, despite overall β-oxidation gene downregulation, could reflect a metabolic adaptation to a Western-style diet high in fructose and fat. Approximately 95% of dietary lipids are triacylglycerols, primarily composed of LCFA,^75^ and ACADVL encodes the main enzyme catalyzing the initial step of LCFA β-oxidation.^76^ Additionally, *CPT1A* expression *–* which facilitates mitochondrial import of LCFAs^77^ – was not decreased in this study potentially reflecting a coordinated response to increased LCFA load.

In addition, higher fasting fructose was associated with reduced *PPARGC1A* expression in our study, which is consistent with the demonstrated mitochondrial dysfunction. Correspondingly, high-fructose feeding in rats impaired mitochondrial biogenesis, indicated by reduced mitochondrial DNA copy number, downregulation of Ppargc1a,^14^ and increased hepatic acetylation of Pgc-1α,^78^ an inactivating modification. Furthermore, reduced mitochondrial biogenesis contributes to mitochondrial dysfunction in MASLD, indicated by lower hepatic expression of *PPARGC1*A and reduced hepatic ETC activity in MASLD patients.^79^ Collectively, these results highlight that the disruption of mitochondrial regulation and function in association with elevated fasting plasma fructose levels can be linked to MASLD, with fasting fructose likely reflecting dietary fructose exposure that contributes to these metabolic alterations. Although fasting fructose was also associated with gut microbial composition, we hypothesize that the association with mitochondrial function primarily reflects a direct hepatic effect of fructose rather than microbiome-mediated mechanisms, given that peripheral fructose concentrations are considered indicative of hepatic fructose exposure.

A significant strength of this study is its focus on associations with fasting fructose levels in humans rather than fructose exposure, providing a clinically relevant perspective and addressing a gap left by predominantly animal-based studies. Furthermore, the integrated approach combining both transcriptional and microbial profiling facilitated examination of host-microbe interactions in the context of fasting plasma fructose and MASLD. Limitations include the small study population consisting exclusively of individuals with morbid obesity, limiting the generalizability of our findings to non-obese individuals or those with BMI <35 kg/m^2,^ and constraining the establishment of clinically and biologically relevant cut-off values for fasting fructose concentrations. Additionally, the absence of small intestinal microbiota data as well as functional microbial analyses, restricts the characterization of gut microbial dynamics in this key site of fructose fermentation and ethanol production. Finally, the cross-sectional design of this study limits the ability to establish causality.

While fasting glucose is a well-established metabolic (glycemic) marker, fasting fructose may provide complementary information due to its distinct hepatic metabolism and effects. Our data challenge the notion that fasting plasma fructose levels merely reflect endogenous production, instead suggesting it primarily indicates dietary exposure based on similarities in associated transcriptional changes to those previously observed in fructose-feeding models. We demonstrated that fasting plasma fructose levels positively correlate with postprandial ethanol excursions in plasma, likely mediated by gut microbial reshaping and impaired host ethanol clearance, as indicated by downregulation of ADH genes. Furthermore, fructose and its catabolic products have been linked to hepatic mitochondrial dysfunction, which we similarly observed with increased fasting plasma fructose. This highlights a key mechanism proposed to contribute to MASLD-associated metabolic disruption. Together, these findings suggest that fasting fructose could be an indicator of hepatic metabolic stress, contributing to the characterization of the metabolic state of the liver. Future longitudinal studies are needed to evaluate whether fasting plasma fructose is a reliable proxy for dietary fructose exposure and can serve as a novel (additional) risk marker for MASLD, while also elucidating the potential causal and mediating mechanisms, including the role of the gut microbiome.

## Data Availability

All data produced in the present study are available upon reasonable request to the authors

## Author information

## Contributions

F.H.M.W., I.A., A.S.M., M.R.M., M.N. conceived the project. A.S.M., A.L. and R.F. managed the prospective BARIA cohort and were involved in the collection of clinical data. J.V. and M.D. were responsible for the histological assessment of liver biopsies. N.F.S. was responsible for the execution and analysis of the *in vitro* experiments. O.A. preprocessed the microbiome data. M.R.M. performed GC-QQQ plasma fructose measurements. F.H.M.W. analyzed all data, except the single-nuclei RNA-seq data which was analyzed by A.S.K.. F.H.M.W., I.A., M.R.M. and M.N. wrote the manuscript, with input from all the authors.

### Acknowledgements

I.A. was funded through an Amsterdam Cardiovascular Sciences MD/PhD grant (2023) and M.R.M. was funded through an Amsterdam Cardiovascular Sciences Postdoctoral Talent grant (2023). A.S.M. is supported by a NWO VENI grant 2023 (09150162310148) and a personal Dutch Digestive Health fund 2024 (WO2423). M.N. is supported by a personal NWO VICI grant 2020 (09150182010020) and an ERC Advanced grant (101141346) on which F.H.M.W. was appointed. The funders of this study had no role in study design, data collection, data analysis, data interpretation, or writing of the manuscript. We would like to thank the Dutch liver pathology panel (M.M. Laclé, Universitair Medisch Centrum Utrecht; P. Drillenburg, Onze Lieve Vrouwe Gasthuis, Amsterdam; A.M. Jonker, Gelre Ziekenhuis, Apeldoorn; A. Farina Sarasqueta, Amsterdam UMC, Amsterdam; C.M. Bronkhorst, Pathologie-DNA, location Jeroen Bosch Ziekenhuis, ’s Hertogenbosch; M. Doukas, Erasmus Medisch Centrum, Rotterdam; N. N. T. Goemaere, Maasstad Ziekenhuis, Rotterdam; I.V. Samarski, Maasstricht Universitair Medisch Centrum, Maasstricht; Arjan van Tilburg, Haga Ziekenhuis, Den Haag).

## Data availability statement

Fecal metagenomics data of the first 100 subjects from the ongoing BARIA cohort have been deposited in the European Nucleotide Archive (ENA PRJEB47902). Liver and jejunum transcriptomics data are available in the European Genome-Phenome Archive (EGAS00001005704). The single-nuclei RNA-sequencing data utilized in this study are publicly available from the original publication^28^ and from the Gene Expression Omnibus (https://www.ncbi.nlm.nih.gov/geo/query/acc.cgi) under accession number GSE244832. Other original data generated in this study are available from the corresponding author upon reasonable request.

