## Supplementary Figures and Tables for "Fasting plasma fructose is associated with postprandial ethanol levels and mitochondrial dysfunction: linking fasting fructose to MASLD"

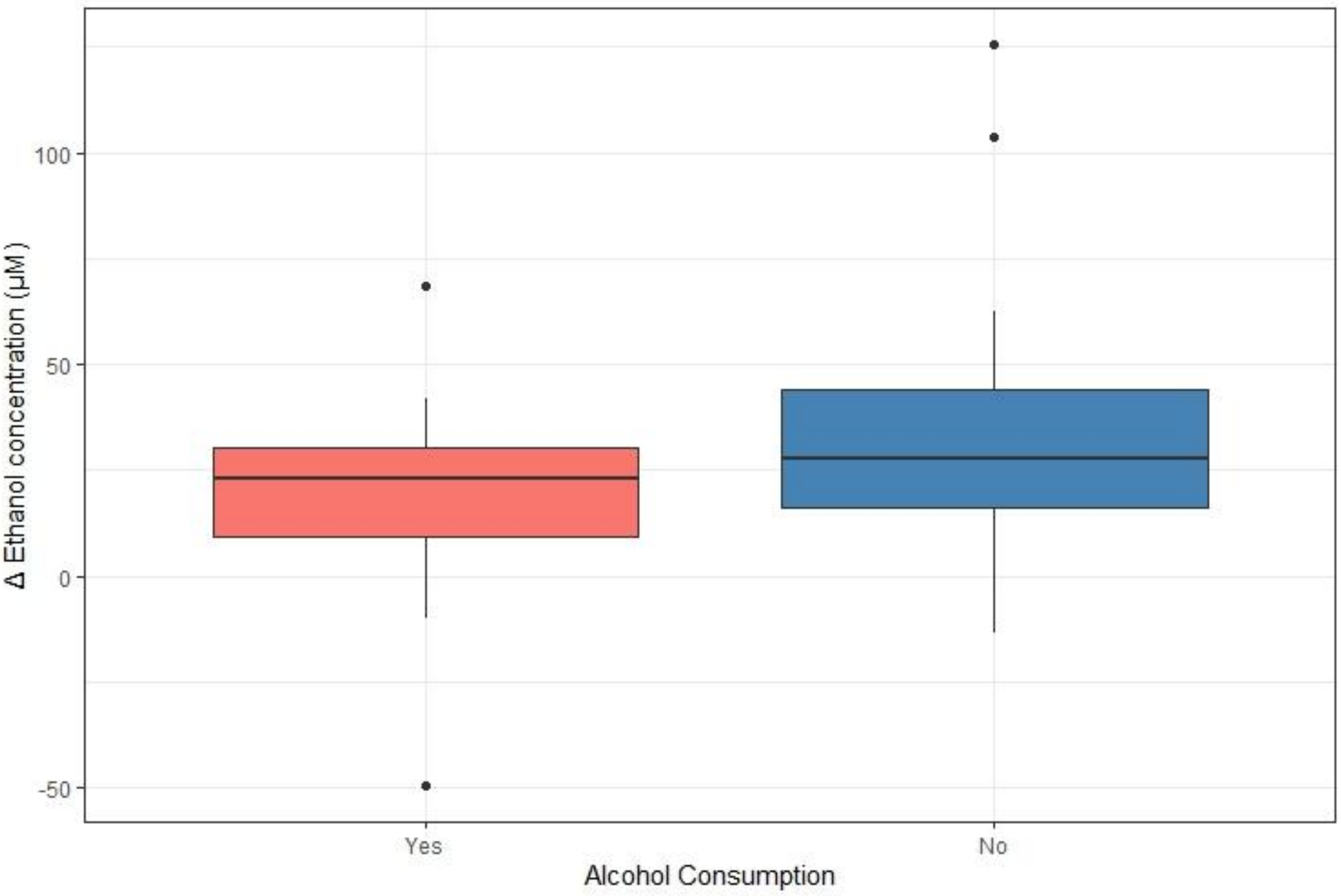

**Supplementary Figure 1. Postprandial plasma ethanol increase stratified by self-reported alcohol consumption.** No significant difference was observed between the two groups (*p*=0.564)

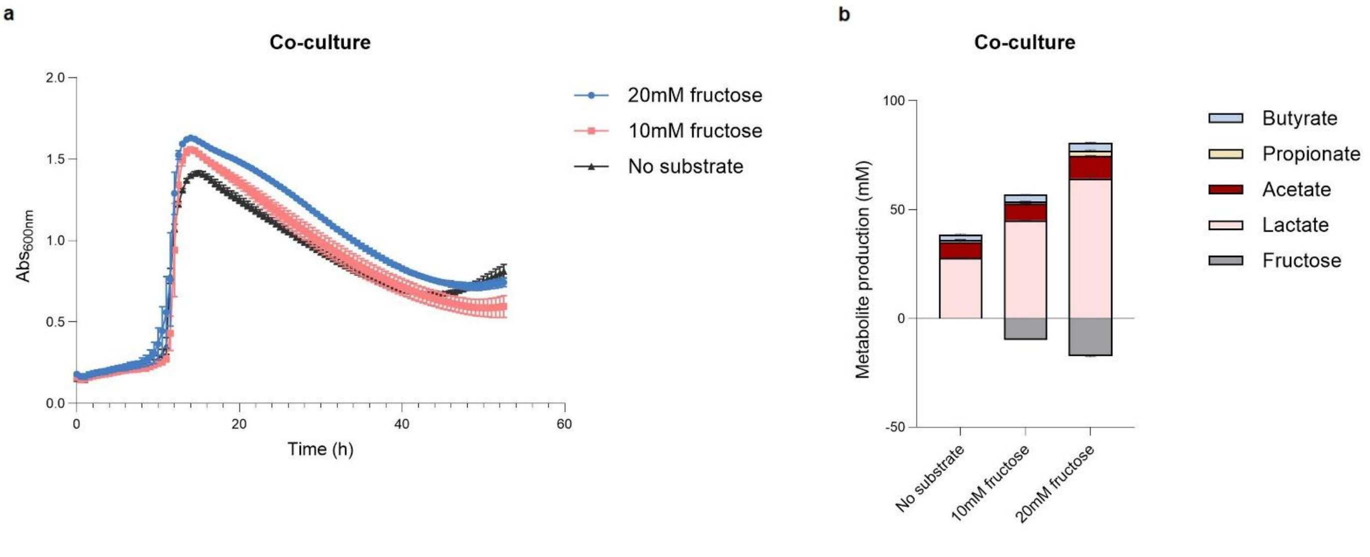

**Supplementary Figure 2. Fructose-dependent growth dynamics and fermentation profiles of *S. parasanguinis* and *G. pamelaeae* in co-culture.** (**a**) Growth kinetics ($Abs_{600nm}$) of the co-culture at 0, 10, and 20 mM fructose. (**b**) Fermentation end-products of the co-culture across fructose concentrations.

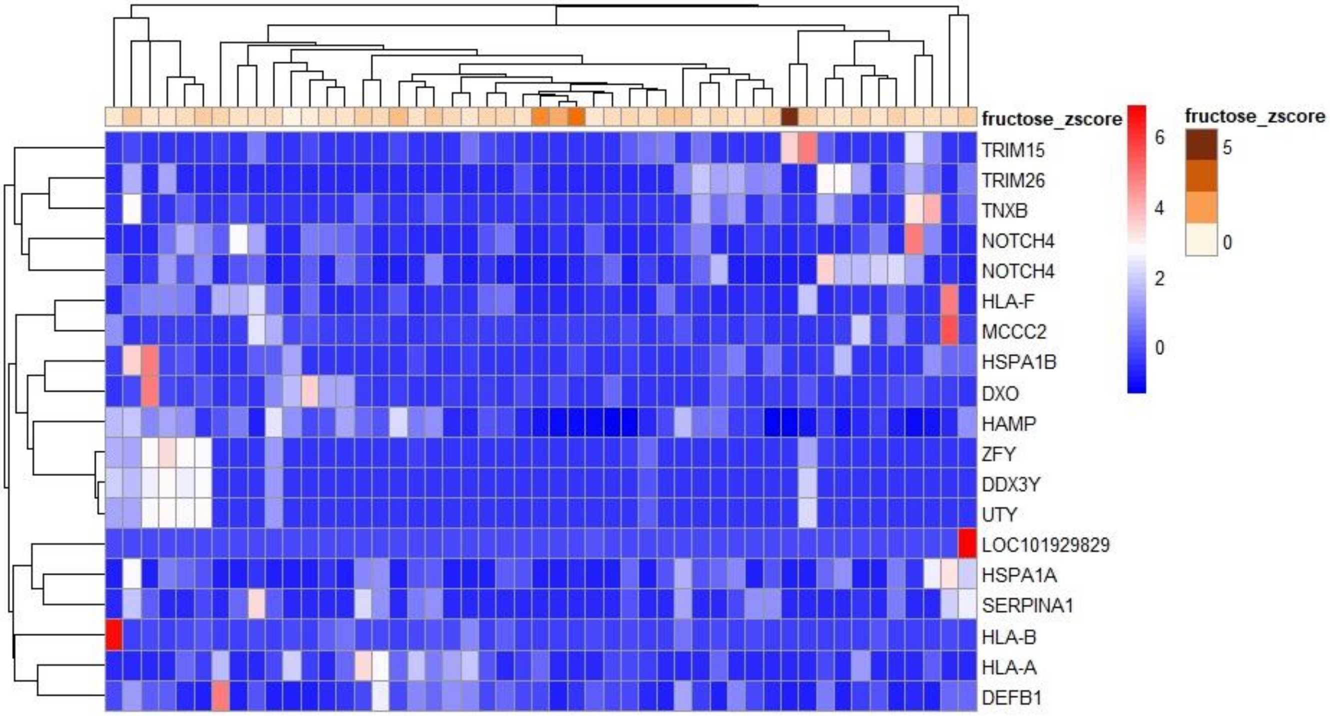

**Supplementary Figure 3. Heatmap of differentially expressed genes.** Heatmap of differentially expressed genes with an absolute Log2FoldChange ≥1. The *fructose_zscore* represents fasting fructose concentrations that were standardized (Z-score normalized) prior to analysis. No clear clustering of gene expression patterns was observed in relation to the fructose levels

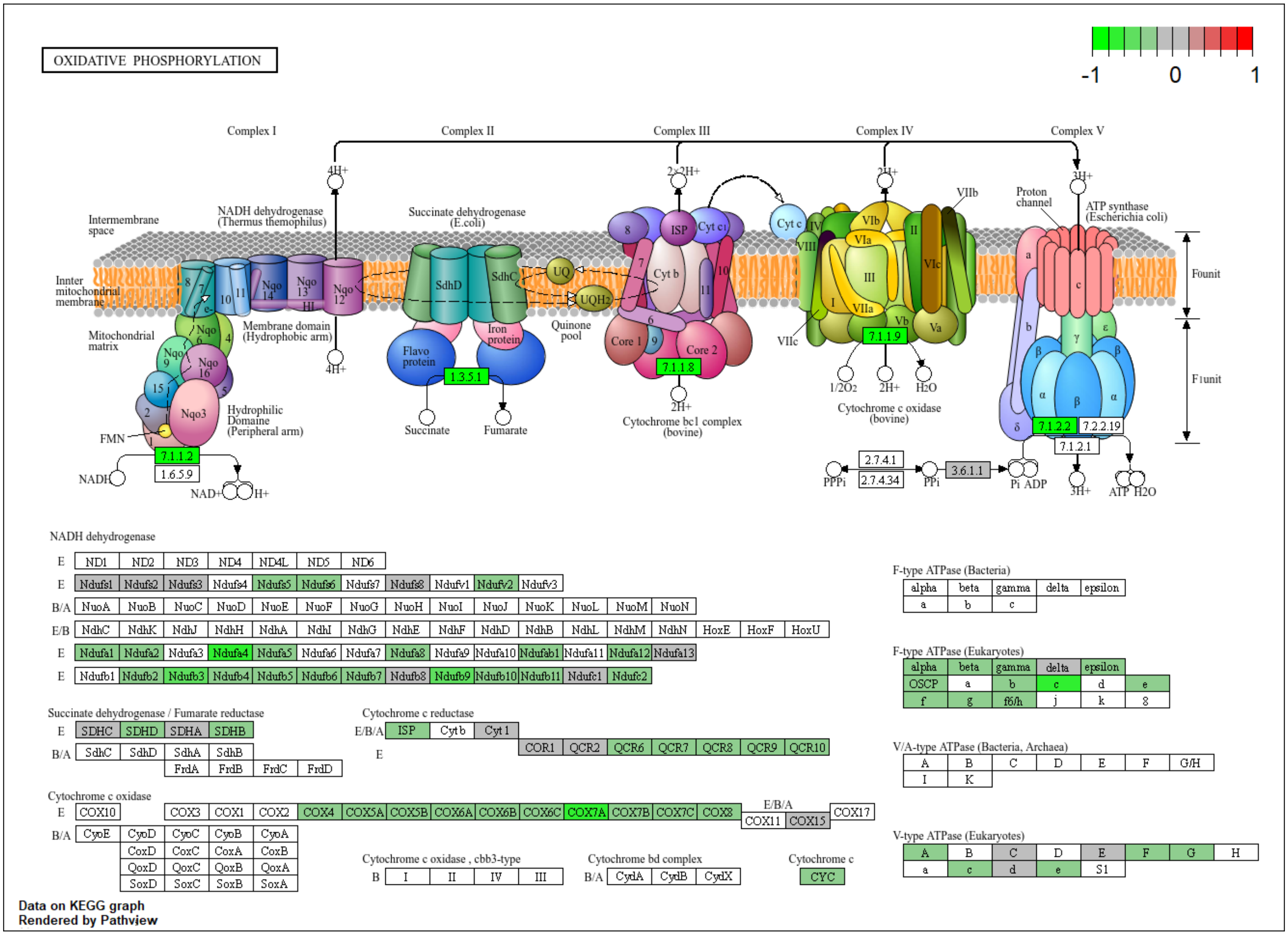

**Supplementary Figure 4. KEGG pathway map of oxidative phosphorylation**

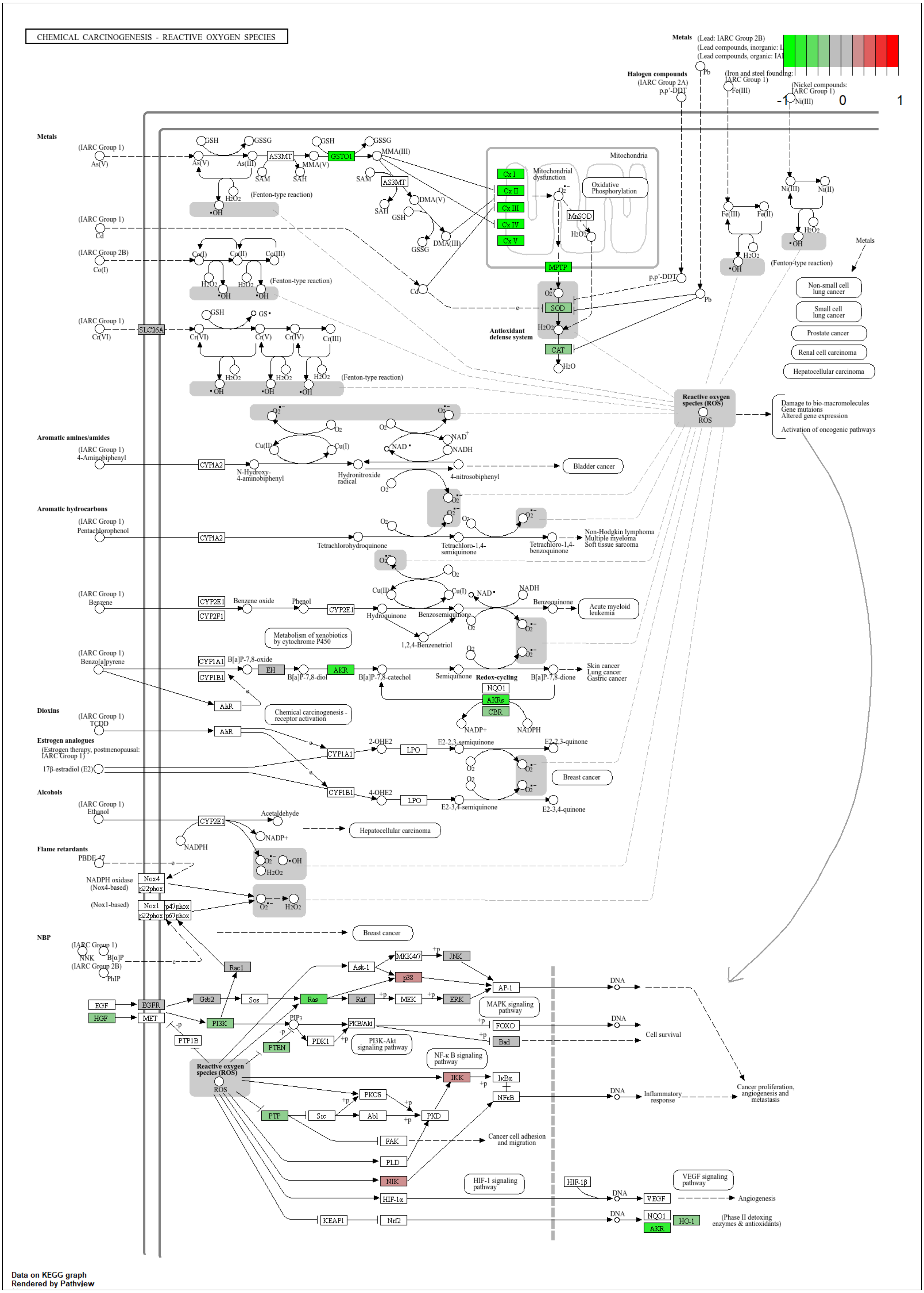

**Supplementary Figure 5. KEGG pathway map of Chemical Carcinogenesis – Reactive Oxygen Species**

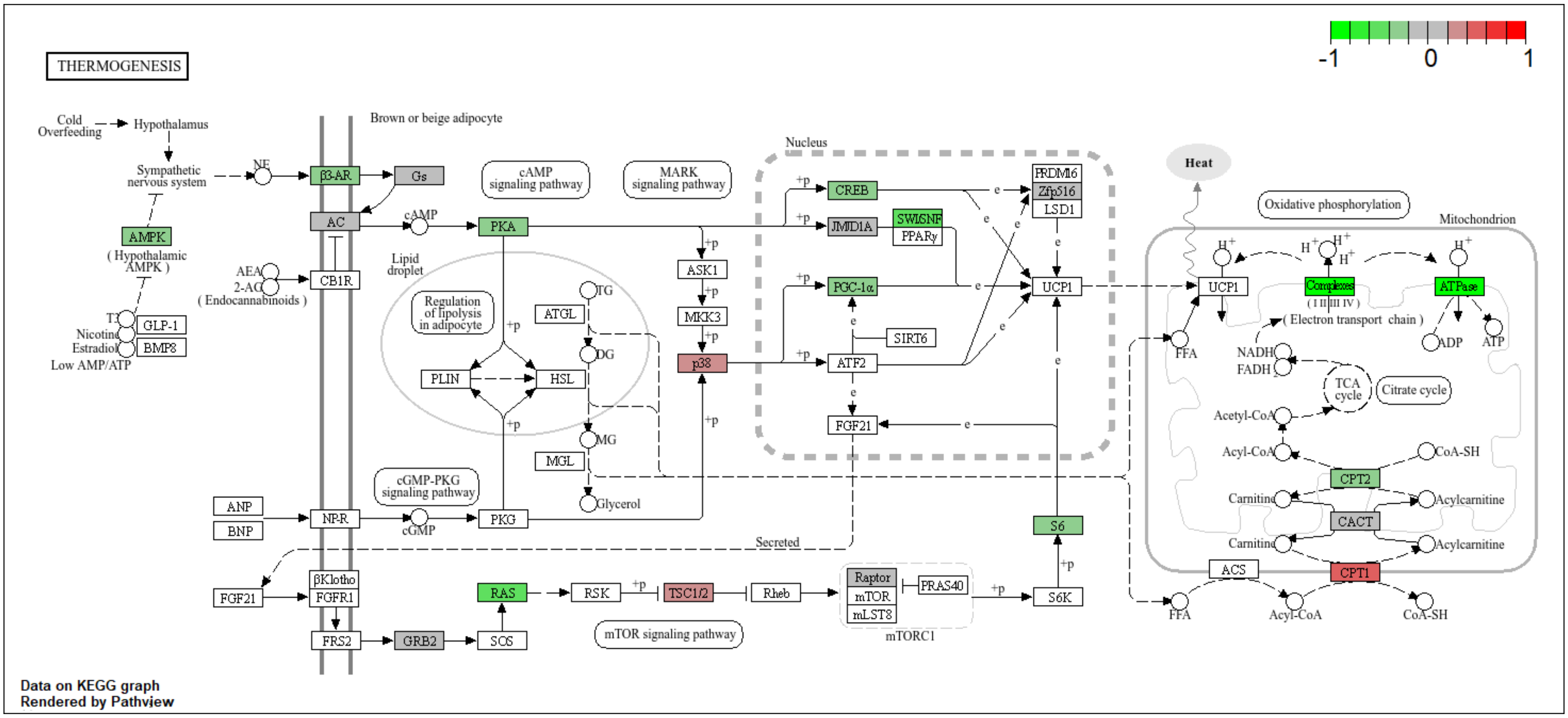

**Supplementary Figure 6. KEGG pathway map of Thermogenesis**

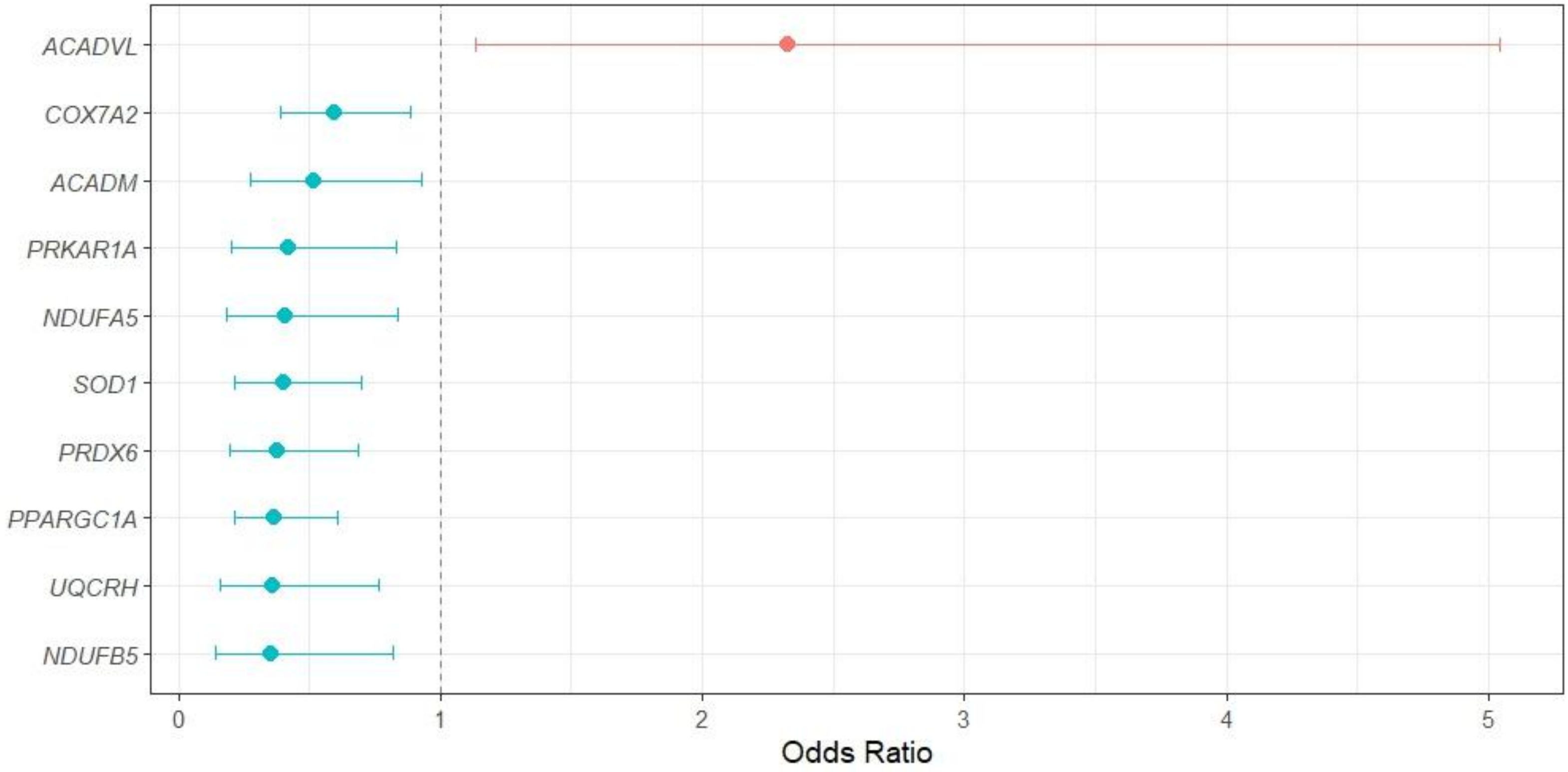

**Supplementary Figure 7. Multivariable logistic regression analyses results associated with increased MASLD risk.** Multivariable logistic regression analyses were performed to evaluate the association between the expression levels of identified DEGs in bulk RNA-seq analysis and MASLD status. Genes that were included in the analyses included the DEGs involved in OXPHOS, β-oxidation and antioxidant defense, as well as regulatory PKA-encoding genes, *CREB1* and *PPARGC1A*. Each gene was tested in a separate model and all models were adjusted for the confounders age and sex. Higher expression of β-oxidation gene *ACADVL* was significantly associated with increased odds of MASLD. Conversely, reduced expression of the β-oxidation gene *ACADM*, OXPHOS-related genes (*NDUFA5*, *NDUFB5*, *UQCRH*, *COX7A2*), antioxidant genes (*SOD1*, *PRDX6*), and regulatory genes (*PRKAR1A*, *PPARGC1A*) were each independently associated with higher MASLD risk. Statistical significance was determined after FDR correction. ACADVL: (OR: 2.33, 95% CI: 1.13-5.04, $p$=0.026), *COX7A2*: (OR: 0.59, 95% CI: 0.39-0.89, $p$=0.027), *ACADM*: (OR: 0.52, 95% CI: 0.28-0.93, $p$=0.050), *PRKAR1A*: (OR: 0.42, 95% CI: 0.20-0.83, $p$=0.028), *NDUFA5*: (OR: 0.41, 95% CI: 0.19-0.84, $p$=0.037), *SOD1*: (OR: 0.40, 95% CI: 0.22-0.70, $p$=0.0047), *PRDX6*: (OR: 0.38, 95% CI: 0.20-0.68, $p$=0.0040), *PPARGC1A*: (OR: 0.37, 95% CI: 0.22-0.61, $p$<0.001), *UQCRH*: (OR: 0.37, 95% CI: 0.16-0.76, $p$=0.014), *NDUFB5*: (OR: 0.35, 95% CI: 0.14-0.82, $p$=0.027).

| ETC complexes | Gene symbol | Description | baseMean | log2FC | *p*-value | *p*adj |
|---|---|---|---|---|---|---|
| **NADH dehydrogenase** | *NDUFA1* | NADH:ubiquinone oxidoreductase subunit A1 | 83.7 | -0.33 | <0.001 | <0.001 |
| | *NDUFA12* | NADH:ubiquinone oxidoreductase subunit A12 | 63.6 | -0.28 | <0.001 | 0.0028 |
| | *NDUFA2* | NADH:ubiquinone oxidoreductase subunit A2 | 205.0 | -0.28 | <0.001 | <0.001 |
| | *NDUFA4* | NDUFA4 mitochondrial complex associated | 464.8 | -0.40 | <0.001 | <0.001 |
| | *NDUFA5* | NADH:ubiquinone oxidoreductase subunit A5 | 332.0 | -0.27 | <0.001 | <0.001 |
| | *NDUFA8* | NADH:ubiquinone oxidoreductase subunit A8 | 69.8 | -0.30 | <0.001 | 0.0049 |
| | *NDUFAB1* | NADH:ubiquinone oxidoreductase subunit AB1 | 191.9 | -0.30 | <0.001 | <0.001 |
| | *NDUFB2* | NADH:ubiquinone oxidoreductase subunit B2 | 252.3 | -0.22 | <0.001 | <0.001 |
| | *NDUFB3* | NADH:ubiquinone oxidoreductase subunit B3 | 86.8 | -0.44 | <0.001 | 0.0015 |
| | *NDUFB4* | NADH:ubiquinone oxidoreductase subunit B4 | 123.8 | -0.38 | <0.001 | <0.001 |
| | *NDUFB5* | NADH:ubiquinone oxidoreductase subunit B5 | 492.4 | -0.23 | <0.001 | 0.0016 |
| | *NDUFB6* | NADH:ubiquinone oxidoreductase subunit B6 | 165.8 | -0.30 | <0.001 | 0.0040 |
| | *NDUFB7* | NADH:ubiquinone oxidoreductase subunit B7 | 182.3 | -0.27 | <0.001 | 0.0014 |
| | *NDUFB9* | NADH:ubiquinone oxidoreductase subunit B9 | 186.5 | -0.41 | <0.001 | <0.001 |
| | *NDUFB10* | NADH:ubiquinone oxidoreductase subunit B10 | 111.0 | -0.32 | <0.001 | <0.001 |
| | *NDUFB11* | NADH:ubiquinone oxidoreductase subunit B11 | 239.9 | -0.31 | <0.001 | <0.001 |
| | *NDUFC2* | NADH:ubiquinone oxidoreductase subunit C2 | 435.6 | -0.28 | <0.001 | <0.001 |
| | *NDUFS5* | NADH:ubiquinone oxidoreductase subunit S5 | 53.8 | -0.24 | <0.001 | 0.0042 |
| | *NDUFS6* | NADH:ubiquinone oxidoreductase subunit S6 | 35.6 | -0.36 | <0.001 | <0.001 |
| | *NDUFV2* | NADH:ubiquinone oxidoreductase core subunit V2 | 672.7 | -0.27 | <0.001 | <0.001 |
| **Succinate dehydrogenase** | *SDHB* | Succinate dehydrogenase complex iron sulfur subunit B | 468.1 | -0.26 | <0.001 | 0.0012 |
| | *SDHD* | Succinate dehydrogenase complex subunit D | 1183.3 | -0.26 | <0.001 | 0.0053 |
| **Cytochrome c reductase** | *UQCR10* | Ubiquinol-cytochrome c reductase, complex III subunit X | 135.1 | -0.40 | <0.001 | <0.001 |
| | *UQCR11* | Ubiquinol-cytochrome c reductase, complex III subunit XI | 231.4 | -0.30 | <0.001 | <0.001 |
| | *UQCRB* | Ubiquinol-cytochrome c reductase binding protein | 310.7 | -0.26 | <0.001 | <0.001 |
| | *UQCRFS1* | Ubiquinol-cytochrome c reductase, Rieske iron-sulfur polypeptide 1 | 375.5 | -0.27 | <0.001 | 0.0019 |
| | *UQCRH* | Ubiquinol-cytochrome c reductase hinge protein | 167.0 | -0.27 | <0.001 | <0.001 |
| | *UQCRQ* | Ubiquinol-cytochrome c reductase complex III subunit VII | 374.3 | -0.34 | <0.001 | <0.001 |
| **Cytochrome c (oxidase)** | *CYCS* | Cytochrome c, somatic | 328.6 | -0.22 | 0.0034 | 0.023 |
| | *COX4I1* | Cytochrome c oxidase subunit 4I1 | 682.0 | -0.26 | <0.001 | <0.001 |
| | *COX5A* | Cytochrome c oxidase subunit 5A | 164.4 | -0.25 | <0.001 | <0.001 |
| | *COX5B* | Cytochrome c oxidase subunit 5B | 449.7 | -0.33 | <0.001 | <0.001 |
| | *COX6Â1* | Cytochrome c oxidase subunit 6A1 | 541.5 | -0.34 | <0.001 | <0.001 |
| | *COX6B1* | Cytochrome c oxidase subunit 6B1 | 161.3 | -0.28 | <0.001 | <0.001 |
| | *COX6C* | Cytochrome c oxidase subunit 6C | 396.6 | -0.30 | <0.001 | <0.001 |
| | *COX7A2* | Cytochrome c oxidase subunit 7A2 | 254.7 | -0.46 | <0.001 | <0.001 |
| | *COX7A2L* | Cytochrome c oxidase subunit 7A2 like | 332.2 | -0.23 | <0.001 | 0.0016 |
| | *COX7B* | Cytochrome c oxidase subunit 7B | 217.7 | -0.33 | <0.001 | <0.001 |
| | *COX7C* | Cytochrome c oxidase subunit 7C | 542.7 | -0.35 | <0.001 | <0.001 |
| | *COX8A* | Cytochrome c oxidase subunit 8A | 339.7 | -0.26 | <0.001 | <0.001 |

| | | | | | | |
|---|---|---|---|---|---|---|
| **F-type ATPase** | *ATP5F1A* | ATP synthase F1 subunit alpha | 2383.0 | -0.20 | <0.001 | 0.0023 |
| | *ATP5F1B* | ATP synthase F1 subunit beta | 2171.4 | -0.25 | <0.001 | 0.0049 |
| | *ATP5F1C* | ATP synthase F1 subunit gamma | 705.6 | -0.24 | <0.001 | <0.001 |
| | *ATP5F1E* | ATP synthase F1 subunit epsilon | 210.6 | -0.29 | <0.001 | <0.001 |
| | *ATP5MC1* | ATP synthase membrane subunit c locus 1 | 153.6 | -0.24 | <0.001 | <0.001 |
| | *ATP5MC2* | ATP synthase membrane subunit c locus 2 | 417.5 | -0.17 | <0.001 | <0.001 |
| | *ATP5MC3* | ATP synthase membrane subunit c locus 3 | 715.6 | -0.31 | <0.001 | <0.001 |
| | *ATP5ME* | ATP synthase membrane subunit e | 34.0 | -0.35 | <0.001 | 0.0039 |
| | *ATP5MF* | ATP synthase membrane subunit f | 146.5 | -0.24 | <0.001 | 0.0016 |
| | *ATP5MG* | ATP synthase membrane subunit g | 145.9 | -0.32 | <0.001 | <0.001 |
| | *ATP5PB* | ATP synthase peripheral stalk-membrane subunit b | 560.2 | -0.28 | <0.001 | 0.0021 |
| | *ATP5PF* | ATP synthase peripheral stalk subunit F6 | 174.3 | -0.37 | <0.001 | <0.001 |
| | *ATP5PO* | ATP synthase peripheral stalk subunit OSCP | 353.5 | -0.24 | <0.001 | 0.0010 |

**Supplementary Table 1. DESeq2 analysis results of downregulated oxidative phosphorylation KEGG pathway genes**. ETC, electron transport chain; log2FC, log2 fold change; *p*adj, adjusted *p*-value; NADH, nicotinamide adenine dinucleotide hydrogen; ATP, adenosine triphosphate

| Gene symbol | Description | baseMean | log2FC | *p*-value | *p*adj |
|---|---|---|---|---|---|
| *CPT1A* | Carnitine palmitoyltransferase 1A | 2232.3 | | NS | NS |
| *CPT2* | Carnitine palmitoyltransferase 2 | 809.1 | -0.21 | <0.001 | 0.0056 |
| *SLC25A20* | Solute carrier family 25 member 20 | 759.5 | -0.19 | 0.0043 | 0.026 |
| *ACADS* | Acyl-CoA dehydrogenase short chain | 1896.2 | -0.09 | 0.024 | 0.081* |
| *ACADM* | Acyl-CoA dehydrogenase medium chain | 1605.1 | -0.23 | <0.001 | 0.0034 |
| *ACADL* | Acyl-CoA dehydrogenase long chain | 546.4 | | NS | NS |
| *ACADVL* | Acyl-CoA dehydrogenase very long chain | 6469.8 | 0.15 | 0.0060 | 0.032 |
| *ECHS1* | Enoyl-CoA hydratase, short chain 1 | 5275.1 | -0.20 | <0.001 | 0.0043 |
| *HADH* | Hydroxyacyl-CoA dehydrogenase | 1475.5 | -0.19 | <0.001 | 0.0054 |
| *HADHA* | Hydroxyacyl-CoA dehydrogenase trifunctional multienzyme complex subunit alpha | 1923.0 | -0.22 | <0.001 | 0.0038 |
| *HADHB* | Hydroxyacyl-CoA dehydrogenase trifunctional multienzyme complex subunit beta | 2359.3 | -0.22 | <0.001 | 0.0060 |
| *ACAA2* | Acetyl-CoA acyltransferase 2 | 4436.6 | -0.42 | <0.001 | <0.001 |

**Supplementary Table 2. DESeq2 analysis results of genes involved in mitochondrial β-oxidation.** *no significant *p*-value after correction for multiple testing. Log2FC, log2 fold change; *p*adj, adjusted *p*-value; PKA, protein kinase A; NS, not significant.

| Antioxidant pathway | Gene symbol | Description | baseMean | log2FC | *p*-value | *p*adj |
|---|---|---|---|---|---|---|
| | SOD1 | Superoxide dismutase 1 | 2222.6 | -0.29 | <0.001 | <0.001 |
| | CAT | Catalase | 7134.1 | -0.20 | <0.001 | <0.001 |
| Glutathione pathway | GCLC | Glutamate-cysteine ligase catalytic subunit | 1663.9 | -0.07 | 0.015 | 0.059* |
| | GCLM | Glutamate-cysteine ligase modifier subunit | 435.5 | -0.18 | <0.001 | 0.0077 |
| | GSS | Glutathione synthetase | 290.9 | | NS | NS |
| | GPX1 | Glutathione peroxidase 1 | 580.5 | -0.26 | <0.001 | 0.0010 |
| | GPX2 | Glutathione peroxidase 2 | 108.3 | -0.34 | 0.045 | 0.12* |
| | GPX3 | Glutathione peroxidase 3 | 1105.9 | -0.21 | <0.001 | 0.0061 |
| | GPX4 | Glutathione peroxidase 4 | 864.0 | -0.14 | <0.001 | 0.0086 |
| | GSTA1 | Glutathione S-transferase alpha 1 | 4007.6 | -0.28 | <0.001 | 0.0014 |
| | GSTA2 | Glutathione S-transferase alpha 2 | 3610.3 | -0.37 | 0.0041 | 0.026 |
| | GSTM1 | Glutathione S-transferase mu 1 | 1077.3 | | NS | NS |
| | GSTT1 | Glutathione S-transferase theta 1 | 537.2 | | NS | NS |
| | MGST1 | Microsomal glutathione S-transferase | 4027.8 | -0.30 | <0.001 | 0.0028 |
| | GLRX | Glutaredoxin | 348.1 | -0.24 | <0.001 | <0.001 |
| | GLRX3 | Glutaredoxin 3 | 347.5 | | NS | NS |
| | GSR | Glutathione-disulfide reductase | 451.8 | -0.08 | 0.040 | 0.11* |
| Thioredoxin system | TXN | Thioredoxin | 356.1 | -0.30 | <0.001 | 0.0032 |
| | TXN2 | Thioredoxin 2 | 323.5 | -0.17 | <0.001 | 0.0013 |
| | TXNRD1 | Thioredoxin reductase 1 | 458.3 | -0.13 | 0.0064 | 0.033 |
| | TXNRD2 | Thioredoxin reductase 2 | 696.2 | | NS | NS |
| Peroxiredoxin system | PRDX1 | Peroxiredoxin 1 | 1318.0 | -0.26 | <0.001 | 0.0017 |
| | PRDX2 | Peroxiredoxin 2 | 572.3 | -0.28 | <0.001 | <0.001 |
| | PRDX3 | Peroxiredoxin 3 | 1846.3 | -0.37 | <0.001 | <0.001 |
| | PRDX4 | Peroxiredoxin 4 | 542.5 | -0.36 | <0.001 | <0.001 |
| | PRDX5 | Peroxiredoxin 5 | 246.0 | -0.28 | <0.001 | <0.001 |
| | PRDX6 | Peroxiredoxin 6 | 2133.9 | -0.23 | <0.001 | 0.0019 |
| Aldo-keto reductase family | AKR1A1 | Aldo-keto reductase family 1 member A1 | 1289.8 | | NS | NS |
| | AKR1C1 | Aldo-keto reductase family 1 member C1 | 2353.3 | -0.17 | 0.011 | 0.046 |
| | AKR1C2 | Aldo-keto reductase family 1 member C2 | 1268.7 | -0.31 | 0.0038 | 0.024 |
| | AKR1C3 | Aldo-keto reductase family 1 member C3 | 803.1 | -0.24 | 0.0053 | 0.030 |
| | AKR1C4 | Aldo-keto reductase family 1 member C4 | 1537.8 | -0.18 | 0.0039 | 0.025 |

**Supplementary Table 3. DESeq2 analysis results of genes involved in antioxidant defense systems**
*****no significant *p*-value after correction for multiple testing. Log2FC, log2 fold change; *p*adj, adjusted *p*-value; NS, not significant.

| | Gene symbol | Description | baseMean | log2FC | *p*-value | *p*adj |
|---|---|---|---|---|---|---|
| **PKA-encoding genes** | | | | | | |
| | *PRKACA* | Protein kinase cAMP-activated catalytic subunit alpha | 498.2 | -0.13 | <0.001 | 0.0025 |
| | *PRKACB* | Protein kinase cAMP-activated catalytic subunit beta | 345.7 | -0.16 | 0.0054 | 0.030 |
| | *PRKAR1A* | Protein kinase cAMP-dependent type 1 regulatory subunit alpha | 1723.0 | -0.23 | <0.001 | 0.0025 |
| | *PRKAR1B* | Protein kinase cAMP-dependent type 1 regulatory subunit beta | 192.1 | | NS | NS |
| | *PRKAR2A* | Protein kinase cAMP-dependent type 2 regulatory subunit alpha | 585.3 | -0.12 | 0.0019 | 0.016 |
| | *PRKAR2B* | Protein kinase cAMP-dependent type 2 regulatory subunit beta | 26.4 | -0.27 | 0.0020 | 0.016 |
| | *CREB1* | cAMP-response element-binding protein 1 | 438.4 | -0.12 | 0.0029 | 0.021 |
| | *PPARGC1A* | Peroxisome proliferator-activated receptor gamma coactivator 1-alpha | 865.8 | -0.22 | 0.0025 | 0.019 |

**Supplementary Table 4. DESeq2 analysis results of PKA-encoding genes, *CREB1* and *PPARGC1A*. ***no significant *p*-value after correction for multiple testing. Log2FC, log2 fold change; *p*adj, adjusted *p*-value; PKA, protein kinase A; NS, not significant.

| Characteristics* | Expanded cohort (*n*=289) |
|---|---|
| **Demographic** | |
| Age (years) | 47.0 [39.0-54.0] |
| Female | 222 (76.8) |
| **Anthropometric** | |
| BMI (kg/m$^2$) | 39.0 [36.6-41.2] |
| Metabolic dysfunction-associated steatotic liver disease | 180 (62.3) |
| Diabetes Mellitus type 2 | 64 (22.1) |
| **Laboratory parameters** | |
| AST (U/L) | 24 [20-29] |
| ALT (U/L) | 31 [23-42] |
| GGT (U/L) | 28 [20-45] |
| HbA1c (mmol/mol) | 5.6 [5.3-6.2] |
| Fasting glucose (mmol/L) | 5.7 [5.3-6.5] |
| Fasting insulin (pmol/L) | 82.5 [54.0-128.0] |
| Triglycerides (mmol/L) | 1.4 [1.1-2.1] |
| Total cholesterol (mmol/L) | 4.8 [4.0-5.5] |
| HDL-cholesterol (mmol/L) | 1.2 [1.0-1.4] |
| LDL-cholesterol (mmol/L) | 3.0 [2.4-3.8] |
| **Histological parameters** | |
| Steatosis grade score *n* (%) | |
| 0 | 109 (37.7) |
| 1 | 123 (42.6) |
| 2 | 39 (13.5) |
| 3 | 18 (6.2) |
| Lobular inflammation score *n* (%) | |
| 0 | 87 (30.1) |
| 1 | 183 (63.3) |
| 2 | 19 (6.6) |
| Hepatocyte ballooning score *n* (%) | |
| 0 | 241 (83.4) |
| 1 | 43 (14.9) |
| 2 | 5 (1.7) |
| Fibrosis stage *n* (%) | |
| 0 | 51 (18.1) |
| 1 | 167 (59.2) |
| 2 | 64 (22.7) |
| 3 | 0 (0) |
| 4 | 0 (0) |

**Supplementary Table 5. Baseline characteristics of the expanded cohort .** *Median [IQR] for continuous variables and *n* (%) for categorical variables
